# Machine learning models for blood pressure phenotypes combining multiple polygenic risk scores

**DOI:** 10.1101/2023.12.13.23299909

**Authors:** Yana Hrytsenko, Benjamin Shea, Michael Elgart, Nuzulul Kurniansyah, Genevieve Lyons, Alanna C. Morrison, April P. Carson, Bernhard Haring, Braxton D. Mitchel, Bruce M. Psaty, Byron C. Jaeger, C Charles Gu, Charles Kooperberg, Daniel Levy, Donald Lloyd-Jones, Eunhee Choi, Jennifer A Brody, Jennifer A Smith, Jerome I. Rotter, Matthew Moll, Myriam Fornage, Noah Simon, Peter Castaldi, Ramon Casanova, Ren-Hua Chung, Robert Kaplan, Ruth J.F. Loos, Sharon L. R. Kardia, Stephen S. Rich, Susan Redline, Tanika Kelly, Timothy O’Connor, Wei Zhao, Wonji Kim, Xiuqing Guo, Yii Der Ida Chen, the Trans-Omics in Precision Medicine Consortium, Tamar Sofer

**Author notes:** Correspondence: Tamar Sofer Center for Life Sciences CLS-934 3 Blackfan St. Boston, MA 02115. These authors equally contributed to the work.

## Abstract

We construct non-linear machine learning (ML) prediction models for systolic and diastolic blood pressure (SBP, DBP) using demographic and clinical variables and polygenic risk scores (PRSs). We developed a two-model ensemble, consisting of a baseline model, where prediction is based on demographic and clinical variables only, and a genetic model, where we also include PRSs. We evaluate the use of a linear versus a non-linear model at both the baseline and the genetic model levels and assess the improvement in performance when incorporating multiple PRSs. We report the ensemble model’s performance as percentage variance explained (PVE) on a held-out test dataset. A non-linear baseline model improved the PVEs from 28.1% to 30.1% (SBP) and 14.3% to 17.4% (DBP) compared with a linear baseline model. Including seven PRSs in the genetic model computed based on the largest available GWAS of SBP/DBP improved the genetic model PVE from 4.8% to 5.1% (SBP) and 4.7% to 5% (DBP) compared to using a single PRS. Adding additional 14 PRSs computed based on two independent GWASs further increased the genetic model PVE to 6.3% (SBP) and 5.7% (DBP). PVE differed across self-reported race/ethnicity groups, with primarily all non-White groups benefitting from the inclusion of additional PRSs.

## Introduction

Polygenic (risk) scores (PRSs) summarize information from many genetic variants across the genome. PRSs are being increasingly developed for risk prediction and for quantifying the inherited predisposition for a given trait or condition. The number/dosage of associated alleles, typically weighted according to effect size estimates from a genome-wide association study (GWAS) for a given phenotype, are summed to produce a PRS for an individual [1]. Commonly, PRS studies involve testing for the association between a PRS and a trait in the target data and estimating its effect. However, PRSs rely on linear relationship between allele counts and the outcome [2] and do not account for potential interactions between SNPs [3] and non-linear associations between genetic variants and the outcome of interest. Linear prediction models that rely on standard PRS therefore usually only explain a fraction of observed genetic variance. Recently, we developed a non-linear machine learning (ML) model that incorporated both individual SNPs and a PRS for predicting predisposition to a certain trait. We showed that it improves Percent Variance Explained (PVE) in an independent test dataset over the standard approach, in which a single PRS is incorporated into a linear prediction model, in a dataset comprising diverse individuals from multiple self-reported race/ethnic groups [4]. However, due to their potential large number, inclusion of individual SNPs may lead to both high computational burden and to model overfitting to the training dataset, where a model performs poorly on a new dataset (i.e., data that were not used in the training dataset). While feature selection tools may be applied to reduce the number of SNPs, e.g., using least absolute shrinkage and selection operator (LASSO [5]; used in Elgart et al. [4]), these tools may be limited by incorporating an assumption of linearity.

Other PRS prediction approaches improve upon the single-PRS models by employing multiple PRSs calculated from several GWASs, also known as “multi-PRS” approaches [6]. The goal of incorporating multiple PRSs in the model is to utilize the discoveries of multiple GWASs (multi-trait, multi-ancestry) and thus boost the model’s performance. Increase in PVE using multi-PRS model compared with the best single-score predictions was reported by [7] in the context of using PRSs of multiple traits. Other studies also reported improvement in association analysis when utilizing multiple PRSs compared with a single PRS for a single trait association, with and without a PRS selection step [7–12]. Overall, studies comparing “single-PRS” approaches and “multi-PRS” approaches showed higher performance of multiple PRS models [6, 13–15]. While some multi-PRS models combine PRSs based on different GWAS, other multi-PRS models construct several PRS from the same GWAS, usually based on multiple p-value (significance) thresholds – typically when using the clump & threshold methodology. With the clump & threshold methodology, PRS construction requires setting a p-value parameter, for which a set of optimal SNPs is selected to calculate the score. However, there is no single optimal p-value threshold that is known *a priori*. Thus, one strategy for multiple clump & threshold PRSs is to construct PRS for several different p-value thresholds and then include all PRSs in the analysis [16]. Coombes et al. [17] proposed to perform a principal component analysis over a set of PRSs calculated for a range of clump & threshold parameter settings and then using the first “PRS-PC” for the association testing. The main motivation behind the method is that the largest amount of variation in the computed PRSs is captured by the first PRS-PC, thus potentially improving discrimination of the phenotype tested. Thus, multi-PRS approaches combining PRSs from multiple GWAS, and approaches combining multiple PRSs from the same GWAS, have been shown to improve PRS models, where the first approach (multiple GWAS PRSs) has been particularly useful for improving PRS models in diverse populations.

Blood pressure (BP) is highly polygenic and, when elevated, is one of the primary risk factors for the development of several cardiovascular diseases such as coronary artery disease and stroke [18, 19]. Nearly half of the adults in the U.S have hypertension [20], with higher prevalence among adults who are Black, compared to other subgroups [21, 22]. PRSs have been developed to predict BP phenotypes across the lifespan [23–26]. In prior work, we showed that combining multiple PRSs based on a few GWAS, from populations of differences ancestral make-ups, improves BP PRS models [13]. Our work also noted that PRS effect sizes and performance vary by strata defined by important clinical BP predictors, such as age groups, biological sex, and obesity. Nonlinear ML models more naturally account for differences in relationship between variables across population subsets. Thus, BP phenotypes may especially benefit from non-linear ML models.

Here, we develop multi-PRS non-linear ML models and assess their association with systolic and diastolic BP. For each of the two BP outcomes, we develop an ensemble machine learning model that makes a BP prediction based on demographic (age, sex, self-reported race/ethnic background, and study center), BMI, and PRSs. The ensemble model consists of two consecutive components – a baseline model and a genetic model. The baseline model predicts a phenotype using the set of covariates without the genetic component, and the genetic model further explains the residuals from the baseline model. We evaluate the use of a linear versus a non-linear model at both the baseline and the genetic model level and assess the improvement in performance when incorporating multiple PRSs based on several GWAS and p-value thresholds. We compare the ensemble model’s performance on the held-out test dataset stratified by self-reported race/ethnic groups. In secondary analyses, we also developed PRSs specific to linkage disequilibrium (LD) regions, referred to as local-PRSs, and evaluated the possibility of using multi-local-PRSs.

## Results

Figure 1 visualizes the study design and major steps in development and assessment of the ensemble model. The ensemble model included two components: a baseline and a genetic model, where we compared multiple constructions of genetic models. We used cross validation to tune model parameters in the training dataset and evaluated the models in the independent test dataset. Models were trained using 70% of the available data from our multi-ethnic dataset and tested in the remaining 30%.

**Figure 1:**
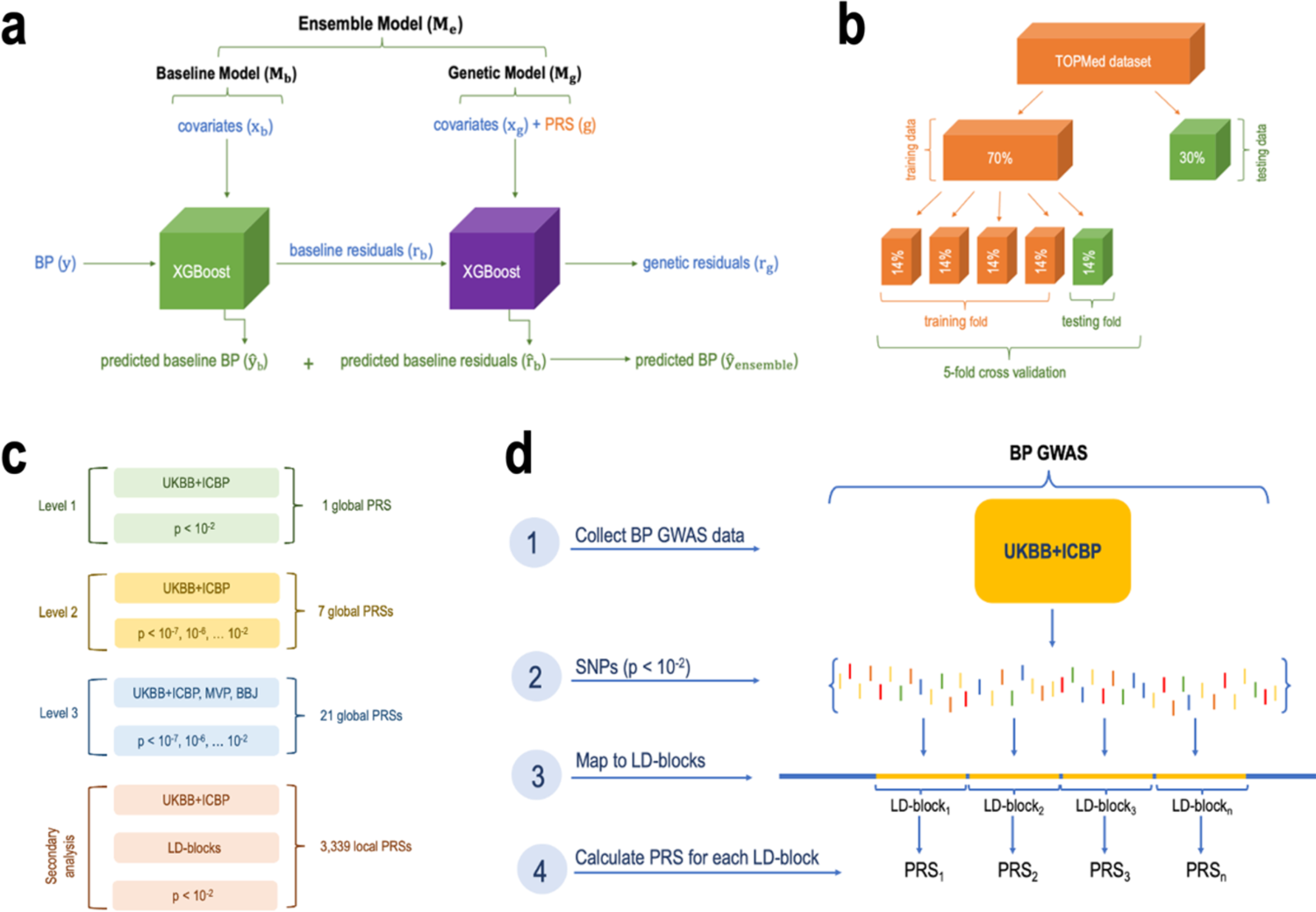
Study design. Panel a: the proposed ensemble model framework. The ensemble is composed of two models. The baseline model, trained on covariates (*X*_b_) only for prediction of SBP and DBP (*y*^_b_). To assess the accuracy of the baseline model we calculated the residuals (baseline residuals *r*_b_) by subtracting the predicted value of SBP/DBP from the actual value of SBP/DBP. The genetic model was trained on a subset of the covariates, and genetic components (global PRSs) for prediction of the baseline model residuals *r*_b_. We measured the accuracy of the genetic model by subtracting predicted genetic residuals *r*^_b_^ from baseline residuals *r*_b_. The overall prediction of BP by the ensemble model is the sum of the predicted baseline BP *y*^ (by the baseline model) and the predicted baseline residuals *r*_b_^ (by the genetic model). The accuracy of the ensemble model was assessed by calculating percent variance explained (PVE) by two models jointly. Panel b: the split of the primary, TOPMed dataset, into training and testing sets followed by the 5-fold cross validation procedure where the training dataset is further split into 5 equal parts with one part designated for testing (repeated 5 times with 1/5 of the training data being designated at random for testing at each iteration). Panel c: increasing levels of genetic models’ complexity where each new model included additional PRSs. Panel d: the process of calculating local PRSs per LD-blocks (secondary analysis). BBJ: BioBank Japan. BP: blood pressure. GWAS: genome wide association study. LD: linkage disequilibrium. Level: model complexity level. MVP: Million Veteran Program. P: p-value threshold. PRS: polygenic risk score. SNPs: single-nucleotide polymorphisms. TOPMed: Trans-Omics for Precision Medicine project. UKBB+ICBP: UK Biobank and International Consortium for Blood Pressure.

### TOPMed participant characteristics

We used a multi-ethnic dataset from the TOPMed consortium (freeze 8 release) to train ML models using PRSs for systolic and diastolic BP (SBP and DBP, respectively). The dataset included 62,295 unrelated participants from fifteen U.S.- and Taiwan-based studies. Individuals self-identified according to categories of race and ethnicity: there were 14,587 Black participants, 30,668 White participants, 4,655 Asian participants, 11,904 Hispanic/Latino participants and 481 participants of “Other” or “Unknown” decent. Descriptions of each of the contributing TOPMed studies are provided in Supplementary Note 1.

The TOPMed dataset was randomly split into a training dataset (70% of the individuals) in which 5-fold cross-validation was used to choose tuning parameters for models, and a held-out test dataset (30% of the individuals) in which the trained models’ predictions were evaluated. Supplementary Table 1 characterizes the training TOPMed dataset. Characteristics of the held-out test TOPMed dataset are provide in Supplementary Table 2. Characteristics broken down by specific TOPMed studies are provided in Supplementary Table 3. In brief, due to the random split to the training and test datasets, both datasets have similar characteristics, including about 49% self-reported White, 23% Black, 19% Hispanic/Latino, 7% Asian individuals and <1% of Other or Unknown race/ethnicity. The mean age is approximately 55 years, 63% of the participants are female, and 59% are hypertensive, with hypertension being more common in self-reported Black individuals and least common in self-reported Asian individuals.

**Table 1:** TOPMed dataset (training and testing sets combined) characteristics aggregated over the studies, stratified by race/ethnic background. Hypertension was defined as SBP≥130, DBP≥ 80, or use of antihypertensive medications. IQR: interquartile range.

| Characteristic | White | Black | Hispanic/Latino | Asian | Other/Unknown |
| --- | --- | --- | --- | --- | --- |
| **N** | 30,668 | 14,587 | 11,904 | 4,655 | 481 |
| **Gender (N (%))** |  |  |  |  |  |
| Female | 20,218 (66%) | 9,165 (63%) | 7,108 (60%) | 2,376 (51%) | 198 (41%) |
| Male | 10,450 (34%) | 5,422 (37%) | 4,796 (40%) | 2,279(49%) | 283 (59%) |
| **Age, years (Median, IQR)** | 60 (50, 69) | 55 (47, 64) | 52 (43, 61) | 48 (40, 57) | 59 (50, 67) |
| **SBP, mmHg (Median, IQR)** | 126 (113, 142) | 133 (119, 150) | 126 (113, 144) | 122 (110, 138) | 134 (116, 152) |
| **DBP, mmHg (Median, IQR)** | 75 (68,83) | 80 (72, 89) | 76 (68, 84) | 75 (68, 85) | 76 (66, 86) |
| **BMI, kg/m<sup>2</sup> (Median, IQR)** | 26.4 (23.5, 30.0) | 29.0 (25.1, 34.0) | 29.0 (26.0, 33.0) | 23.9 (21.8, 26.2) | 27.3 (24.3, 31.5) |
| **Hypertensive (N (%))** | 17,274 (56%) | 10,103 (69%) | 6,733 (57%) | 2,381 (51%) | 323 (67%) |

### Non-linear ML models improve modelling of covariate effects compared to linear models

For each BP outcome, we trained ensemble models using the TOPMed training dataset. The first model in the ensemble, referred to as the baseline model, included the covariates age, sex, BMI, race/ethnic background, and study center, where the latter is a dataset-specific variable. Figure 2 visualizes the phenotypic independent test dataset PVE obtained from the baseline model when fitted using a non-linear ML model (a gradient boosting trees model fitted using the XGBoost package) and using a linear model (see Supplementary Table 4 for baseline model complete results). The ML model had higher PVE for both SBP and DBP, and when evaluated over the complete dataset and by groups defined by self-reported race/ethnic background.

**Figure 2:**
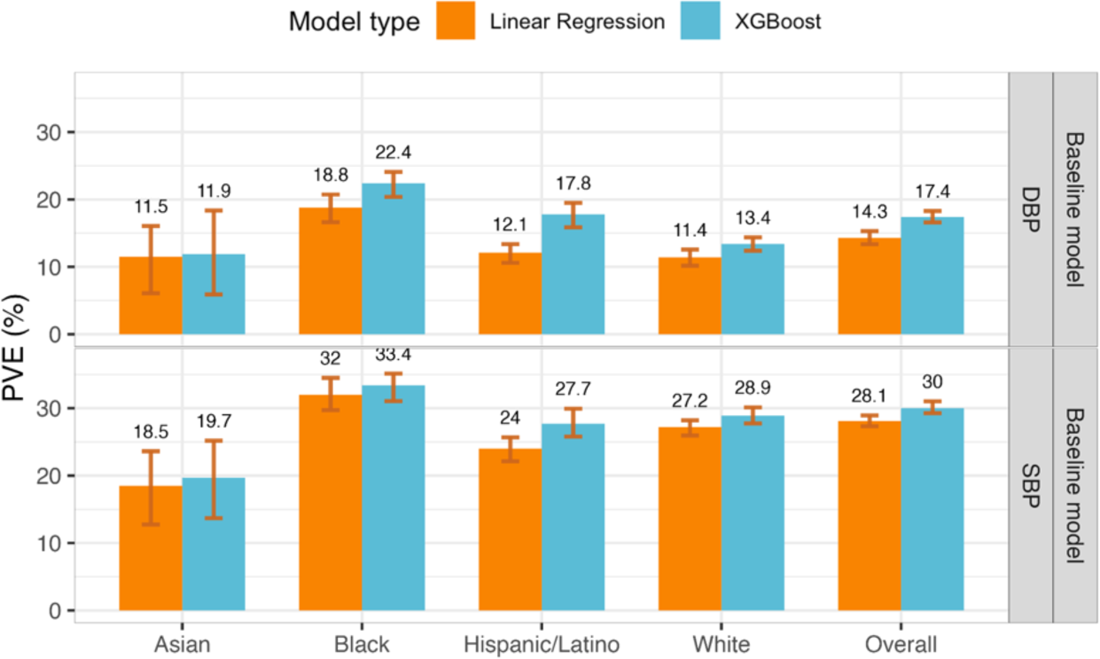
Estimated phenotypic PVE of baseline models fitted using non-linear ML and linear models. Estimated PVEs in the TOPMed test dataset for baseline model performance for prediction of SBP and DBP in the overall test dataset and stratified by self-reported race/ethnicity (White N = 10,877, Hispanic/Latino N = 3,831, Black N = 3,657, Asian N = 403 for DBP; White N = 10,823, Hispanic/Latino N = 3,877, Black N = 3,674, Asian N = 374 for SBP). The visualized 95% confidence intervals were computed as the 2.5% and 97.5% percentiles of the bootstrap distribution of the PVEs estimated over the test dataset. PVE: Percent variance explained. TOPMed: Trans-Omics in Precision Medicine project. SBP: systolic blood pressure. DBP: diastolic blood pressure.

Therefore, we proceeded with ensemble models with non-linear ML baseline model. Complete results, including from cross-validated PVE in the training datasets, are provided in Supplementary Table 5. Surprisingly, we observed slightly lower performance in models trained using global SBP/DBP PRSs developed using Bayesian approach, PRS-CSx (complete results are reported in the Supplementary Table 6). The hyperparameters for the XGBoost model and their values selected after tuning are listed in Supplementary Table 7. For the baseline ML model, the phenotypic PVE was higher for SBP prediction (30% PVE in the race/ethnicity combined dataset) than for DBP (17.4% phenotypic PVE), as reported in other PRS studies of the two phenotypes. When tested on the testing set stratified by race/ethnicity, the PVE was highest in the group of Black individuals, for both SBP and DBP (33.4% and 22.4%, respectively). PVE was lowest in the group of Asian individuals (19.7% and 11.9% for SBP and DBP, respectively).

In Supplementary Figure 1, we also report results from a secondary analysis comparing baseline models with and without inclusion of genetic PCs, demonstrating that PCs inclusion does not improve the baseline model PVE, perhaps because the PRSs already capture BP-related information that related to population structure and that is typically captured by genetic PCs. Thus, and given that use of PCs challenges the transferability of prediction models between datasets, we did not include them.

### Using multiple PRSs from the same and from multiple GWASs improves model performance in Asian, Black, and Hispanic participants

The second part of the ensemble model is the genetic model, which included covariates that can be used across datasets, including age, sex, BMI, self-reported race/ethnic background, and PRS measures. The ensemble model used baseline model (because it performed better than linear regression), and genetic models fitted using either non-linear ML or using conventional linear regression. Further, genetic models were of increasing complexity where we included one or more PRSs according to the following logic: model complexity 1 included a single PRS based on the clump & threshold methodology using p-value threshold of 0.01 and using summary statistics from the BP GWAS of the UK Biobank and the International Consortium of Blood Pressure (UKBB+ICBP), which yielded the most powerful single-GWAS PRSs in the a past paper developing BP PRSs [27]. Model complexity 2 included 7 PRSs based on the clump & threshold methodology each using a different p-value threshold for SNP inclusion, and using the same UKBB+ICBP GWAS. Model complexity 3 included 21 PRSs, 7 PRSs from each of the UKBB+ICBP, Million Veteran Program (MVP), and Biobank Japan (BBJ).

Supplementary Table 5 reports the complete performance results as attained PVEs from the genetic models (PVEs of predicting residuals from the baseline model) and ensemble models (PVEs of predicting the raw trait) estimated in cross validation on the training dataset, and from the independent test dataset. Genetic models fitted using the non-linear ML approach tended to have similar or better performance than genetic model fitted using linear regression (see Supplementary Figure 2). Two exceptions were linear model performed better are prediction of DBP in Black individuals (2.2% PVE by model complexity level 3 linear regression compared with 1.7% PVE of the ML model) and prediction of SBP in Hispanic/Latino individuals (5.7% versus 4.6% in linear regression versus ML level 3 models). We therefore here focus on the genetic models (ML fitted using XGBoost).

Figure 3 visualizes the genetic model performance, measured by PVEs in prediction of residuals from the baseline model, and the ensemble model performance, measured by PVEs at the phenotypic level. Genetic model performance improved with the addition of PRSs, with improvement being large for the non-White groups, and low and almost not existing, for the group of self-reported White individuals. This is likely because the self-reported White individuals are mostly of European genetic ancestry, closely matching the genetic ancestry of the population participating in the UKBB+ICBP GWAS. Concretely, genetic model performance in the White group were 7.7%, 7.6%, and 7.8% for the three increasing complexity levels for SBP, and 7.3%, 6.7%, and 7.2% for DBP. In other individuals the improvement was strongly apparent. In Hispanic/Latino individuals and prediction of SBP, the PVEs were 0.9%, 2%, and 4.6%, and for DBP they were 1.5%, 2.7%, and 5%. In the Asian group the SBP PVEs were 1.6%, 2.4%, and 3.8%, and for DBP they were 0.9%, 2.9%, and 3.4%. Finally, Black individuals had the lowest genetic model performance with PVEs by complexity for SBP being 0.7%, 1.1%, and 2.6%, and for DBP 0.8%, 0.3%, and 1.7%. Therefore, including PRSs based on non-European GWAS summary statistics substantially contributed to the model performance in non-White individuals but only to a small extent, and only for DBP, for White individuals.

**Figure 3:**
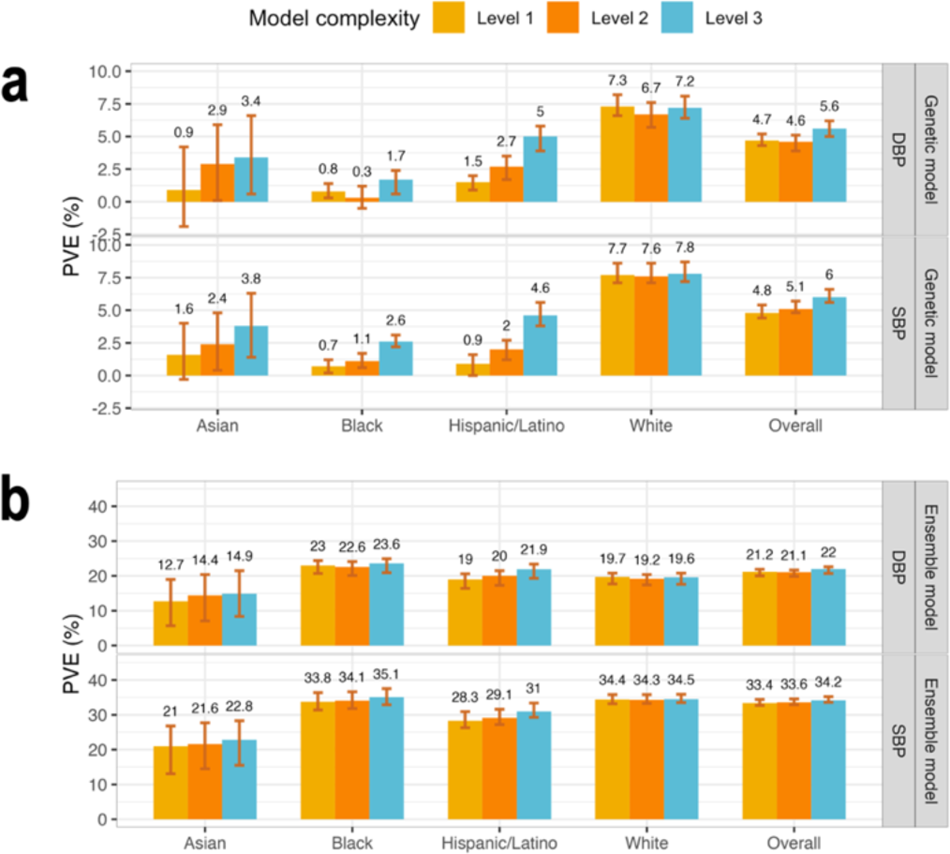
Comparison of genetic and ensemble model performance in TOPMed test dataset. Panel a: Estimated PVEs in the TOPMed test dataset obtained by genetic models incorporating one or more PRSs according to the three complexity levels. Level 1: a single PRS based on the UKB+ICBP GWAS. Level 2: PRSs based on the UKB+ICBP GWAS based on seven p-value thresholds. Level 3: 21 PRSs, 7 PRSs based on each of the UKB+ICBP, MVP, and BBJ GWAS. PVE is reported for predicting residuals from the baseline model, where the baseline model was a non-linear ML model and only used non-genetic covariates. Panel b: Estimated PVEs in the TOPMed test dataset for ensemble model at the raw phenotypic level. PVEs are reported for models of SBP and DBP, in the overall test dataset and stratified by self-reported race/ethnicity (White N = 10,877, Hispanic/Latino N = 3,831, Black N = 3,657, Asian N = 403 for DBP; White N = 10,823, Hispanic/Latino N = 3,877, Black N = 3,674, Asian N = 374 for SBP). The visualized 95% confidence intervals were computed as the 2.5% and 97.5% percentiles of the bootstrap distribution of the PVEs estimated over the test dataset. PVE: Percent variance explained. TOPMed: Trans-Omics in Precision Medicine project. SBP: systolic blood pressure. DBP: diastolic blood pressure. PRS: polygenic risk score.

At the phenotypic, ensemble-model level, the improvement in PVE achieved by increasing the complexity of the genetic models is less impressive, because the covariates explained the lion’s share of the phenotypic variance. Interestingly, while the genetic model had the highest level of PVE in the group of White individuals, the ensemble model, as a whole, had the highest PVE in the group of Black individuals. This was true for both SBP and DBP, and is already seen when using model complexity level 2 (multiple PRSs based only on the UKB+ICBP GWAS). Supplementary Table 6 provides the tuning parameters selected by the cross validation for each of the models.

### Secondary analysis: using local PRSs in the genetic model

In secondary analysis, we developed a new model that included local PRSs (Figure 4a), constructed based on the UKBB+ICBP GWAS, with each local PRS being based on summary statistics restricted to an LD region, with regions defined according to European populations. These PRSs used the clump & threshold methodology with p-value threshold <0.01. Because gradient boosting trees models tend to overfit to the training dataset when many features are included, we extended the development of the ensemble model to include a feature selection step using LASSO penalized regression, as described in Figure 4a.

**Figure 4:**
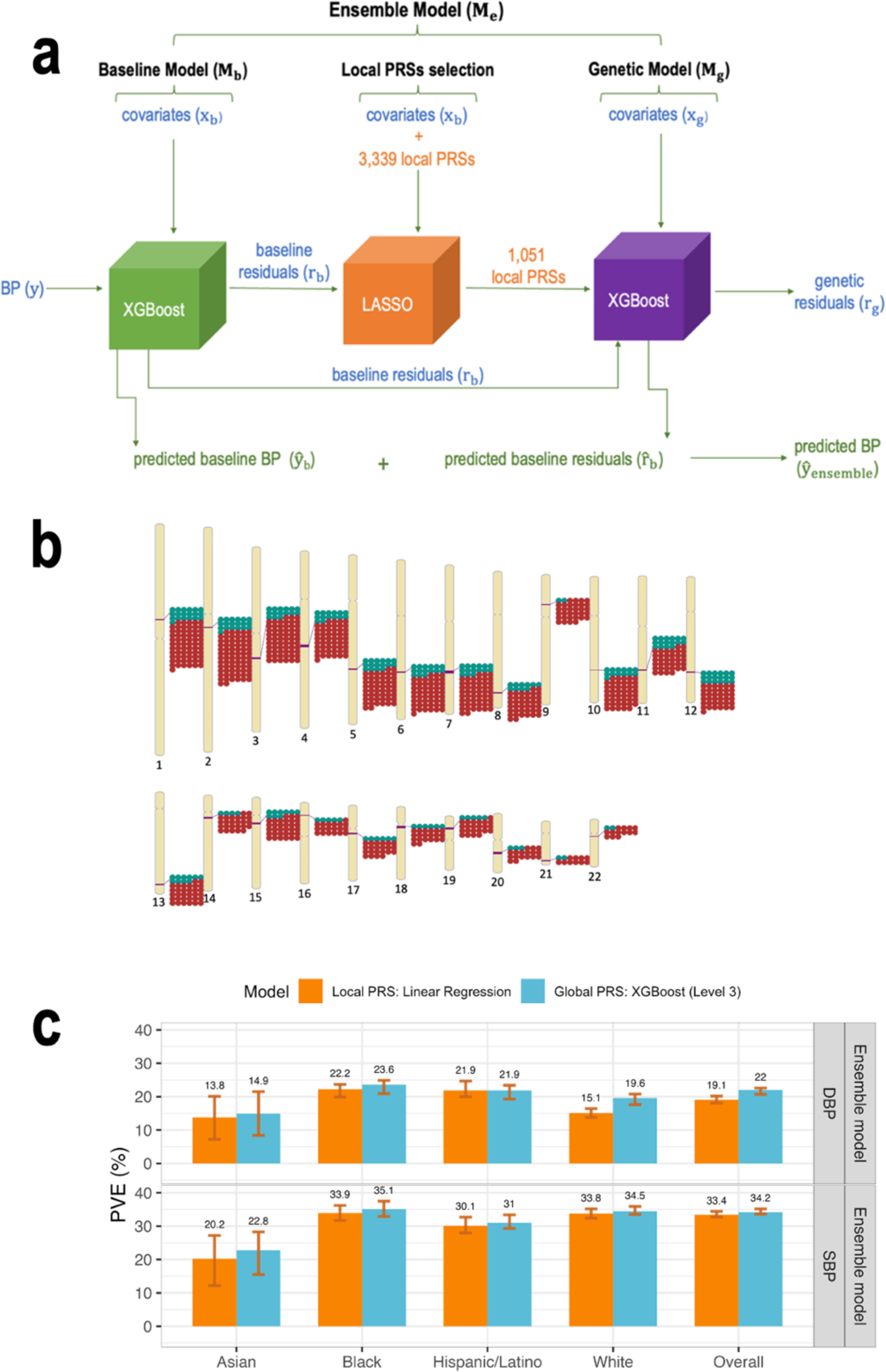
Integration of LASSO feature selection tool into the ensemble model workflow Panel a: The workflow of the ensemble model with the integration of the LASSO variable selection tool. To include local PRSs in the ensemble model while attempting to avoid overfitting, we added a LASSO selection step to the ensemble model development. As visualized, the residuals of the baseline model were used as the outcome in LASSO penalized regression with the local PRSs as features. LASSO substantially reduced the number of local PRSs (to 827 for SBP and 224 for DBP). The local PRSs selected by LASSO were then used as an input into the genetic model for prediction of the baseline residuals (*r*_b_^). Panel b: Genomic locations of local PRSs, calculated over predefined LD-regions, selected by LASSO for SBP and DBP. Panel c: Comparison between the estimated PVE in the TOPMed test dataset for ensemble model Level 3 using global PRSs and the ensemble model using Linear regression and local PRSs. PVEs are reported for models of SBP and DBP, in the overall test dataset and stratified by self-reported race/ethnicity (White N = 10,877, Hispanic/Latino N = 3,831, Black N = 3,657, Asian N = 403 for DBP; White N = 10,823, Hispanic/Latino N = 3,877, Black N = 3,674, Asian N = 374 for SBP). The visualized 95% confidence intervals were computed as the 2.5% and 97.5% percentiles of the bootstrap distribution of the PVEs estimated over the test dataset. BP: blood pressure. DBP: diastolic blood pressure. LASSO: least absolute shrinkage and selection operator. PRS: polygenic risk score. SBP: systolic blood pressure.

As before, we considered either non-linear ML or linear regression genetic models (using the LASSO-selected local PRSs). For comparison, we additionally used an ensemble model that used the fitted LASSO model itself as the genetic model. Note that the difference between the ensemble model with linear regression and with LASSO genetic model is that the linear regression model re-evaluated the coefficients of the local PRSs, while the ensemble model with the LASSO genetic model used the ℓ_!_-penalized coefficients from the LASSO operation.

We evaluated the SBP and DBP local PRS ensemble models’ PVE. LASSO selected a subset of local PRSs which are most salient in model’s prediction (i.e., with the lowest cost function). Specifically, out of 1,670 local PRSs for SBP LASSO selected 827, and it selected 224 of the 1,669 local PRSs for DBP. As shown in Figure 4b, the selected local PRSs for SBP and DBP are concentrated in the same genomic regions, and, more generally, selected local PRSs are clustered in specific regions.

Supplementary Figure 3 provides the PVE of the genetic and ensemble models implemented with the three models (non-linear ML, linear regression, and LASSO). Across self-reported race/ethnic groups the results varied and there is no one method that is always superior to others. Notably, non-linear ML genetic model is almost never the best model. Figure 4c compares the performance of the global PRS model (level 3) with the local PRS model that used a linear regression genetic model (because it tended to perform better than other genetic model specifications). The local PRS genetic model sometimes performed equally well compared to the global PRS model, despite using less information (local compared to genome-wide PRS).

As the local PRSs are based on the UKBB+ICBP GWAS and were generated using the 10^-2^ p-value threshold, it is interesting to compare the genetic model that used them to the genetic model with a single UKBB+ICBP PRS (model complexity level 1). Considering the combined TOPMed test dataset, for SBP, the genetic model using linear regression and local PRSs has better PVE compared to the linear regression genetic model with one global PRS, (PVE=4.8% versus 4.4%). However, this was not true for DBP (linear regression local PRS model PVE =2.6% versus 4.4% for global, single PRS linear regression). The ML global PRS model with complexity level 1 performed the same (for SBP) or better (DBP) than linear regression local PRS models.

Surprisingly, when focusing on Hispanic/Latino individuals, local PRS models performed substantially better than global PRS complexity level 1 models: genetic local PRS model PVEs ranged from 1.6% to 5.7% (across methods and BP traits), while global PRS level 1 model PVEs were at most 1.5%. Supplementary Table 8 reports complete performance results (attained PVEs) from the genetic models (PVEs of predicting residuals from the baseline model) and ensemble models (PVEs of predicting the raw trait) estimated in cross validation on the training dataset, and from the independent test dataset using local PRS in TOPMed dataset.

### Challenges of model generalization to clinical data

Figure 5 visualizes workflow with the application of the model trained on the TOPMed dataset to the MGB Biobank data. For calibration, we first trained a baseline model on the MGB Biobank data (covariates only) and calculated residuals. Next, we applied the genetic model trained on the TOPMed dataset to predict the residuals for the MGB Biobank dataset. Because MGB Biobank data is enriched with data related to hospital-based visits, we restricted the analysis to individuals with no antihypertensive related codes in the two years prior to data extraction (5/25/2021-5/25/2023), and only participants with data from this time period were used. There were 9,494 individuals meeting these inclusion criteria. Of these 9,494 were White, 412 Black, and 200 Asian. The mean age was 59, there are 63% female individuals. Supplementary Table 9 characterizes the study population.

**Figure 5:**
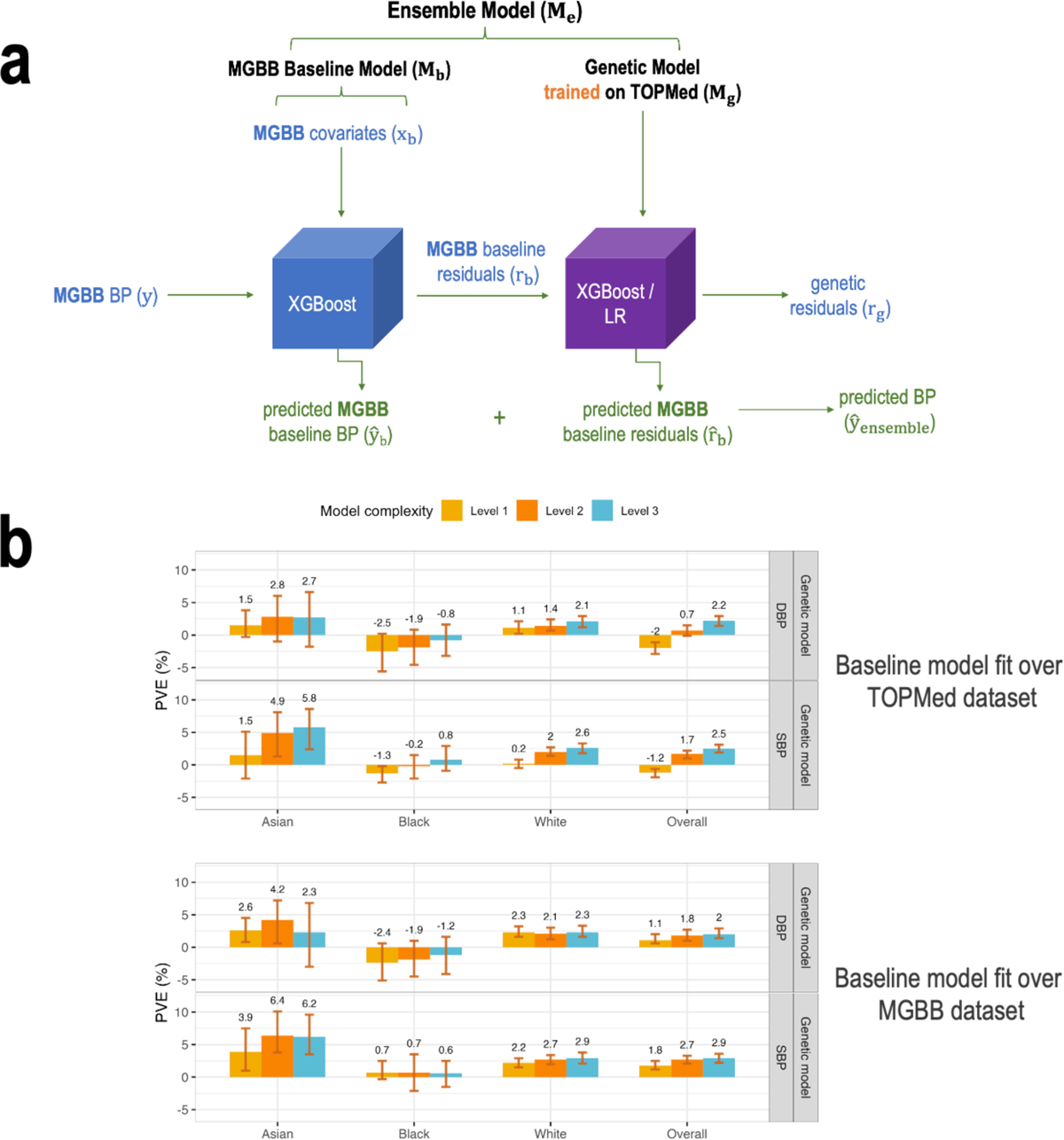
Application of the model trained on the TOPMed dataset to the MGB Biobank data. Panel a: The figure visualizes the workflow of the Ensemble model with the baseline being trained on the MGB Biobank dataset and application of the genetic models, trained on the TOPMed data, incorporating one or more PRSs according to the three model complexity levels. Level 1: a single PRS based on the UKB+ICBP GWAS. Level 2: seven PRSs based on the UKB+ICBP GWAS, difference p-value thresholds. Level 3: 21 PRSs, 7 PRSs based on each of the UKB+ICBP, MVP, and BBJ GWAS. Panel b: Estimated PVE in the MGBB test dataset for XGBoost genetic models fitted on the TOPMed dataset of three levels of complexity with baseline model fitted using TOPMed baseline model weights (top) and using MGBB baseline model weights (bottom). PVEs are shown for the performance in prediction of the second order of residuals for SBP and DBP phenotypes in the overall test dataset and stratified by race/ethnicity (White N = 7,985, Black N = 412, Asian N = 200). BP: blood pressure. MGBB: Mass General Brigham Biobank. PRS: polygenic risk score. TOPMed: Trans-Omics in Precision Medicine project.

Despite strict inclusion criteria, models performed less well on the MGB Biobank dataset. The baseline model was fit in MGB Biobank, for calibration purposes, i.e., to account for differences in populations between the TOPMed and MGB Biobank datasets. Note that the MGB Biobank baseline model was fit without a training-testing approach. We compared the approach of calibrating the baseline model to a new dataset. Figure 5b demonstrates the PVE of the TOPMed-trained genetic models with the baseline model being either TOPMed-trained or MGB-trained. One can see that the calibration approach worked well, in that genetic model performance was usually better when applied over the MGB-trained baseline model.

Supplementary Figure 4 provides the cross-validated PVEs from the baseline ML model trained on the MGB Biobank. The cross-validated PVEs are often lower than the TOPMed test dataset PVEs (Figure 2), other than in the Asian group, where the PVEs are similar. Supplementary Figure 5 provides the PVEs from both the TOPMed-trained genetic model applied on MGB residuals (as in Figure 5), and from the full calibrated ensemble (MGB baseline model + TOPMed-trained genetic model).

## Discussion

The objectives of this study were (a) to develop an ML-based prediction model for BP phenotypes that can more accurately predict BP phenotypes compared to standard linear regression model that includes a single genome-wide PRS model, (b) assess the usefulness of including multiple PRSs in the association model as an alternative to the inclusion of individual SNPs, as the latter option risks overfitting, and (c) examine an approach for calibrating an ML model to a new dataset. To accomplish these goals, we constructed an ensemble model, successively training first on a set of commonly available covariates (demographic variables and BMI), and then on genetic components (PRSs), to directly evaluate the usefulness of an ML compared with a linear regression-based models at both steps. We further developed and compared polygenic models that employ a few levels of PRS inclusion: a single genome-wide PRS constructed based on a single powerful GWAS, multiple genome-wide PRSs constructed based on the same GWAS, and multiple genome-wide PRSs from multiple independent GWASs.

We examined the improvement that such models confer to BP models in groups defined by self-reported race/ethnicity. We also explored the construction of genetic models that use local PRSs, constructed based on LD regions, and, finally, studied the potential to apply the constructed models, and to calibrate model, to an EHR-based dataset using the MGB Biobank dataset.

We quantified model performance using PVEs computed on both the overall, phenotypic level, and at the level of the residuals from the baseline (non-genetic) model. Consistently with other studies, higher PVE was achieved for SBP compared with DBP. For both phenotypes, it was clearly beneficial to use an ML model at the baseline level (consistent with other studies [28]), while an ML genetic model performed better than a linear regression genetic model primarily in self-reported White individuals. Regardless of type of model, using multiple PRSs, including those constructed based on the same GWAS using different p-value thresholds, improved model PVE especially in non-White individuals. In self-reported White individuals using multiple PRSs had relatively little improvement in test dataset PVE. This finding suggests that (a) well-powered GWAS from European-ancestry populations is sufficient to construct good PRS in White individuals (who are primarily of European genetic ancestries); this may be true for individuals of other genetic ancestries, yet data is not yet available to prove it, and (b) GWAS from non-European ancestry populations are useful for PRS in non-White individuals, as is known, and (c) PRSs based on European ancestry GWAS are indeed useful in non-White individuals, and flexibly allowing for potentially different varying association effects of PRSs, each based on different p-value thresholds, is useful. Note that the usefulness of incorporating multiple PRSs based on the same GWAS is immediately apparent. On the face of it, one strong PRS based on a high p-value threshold should be sufficient. The usefulness of multiple PRSs based on the same GWAS resonates with the variability in the contribution of different genetic variants, beyond what is captured in GWAS. We hypothesized the local PRSs may capture this variability better, presumably because different genomic regions have potentially different associations with BP in different populations due to interactions with environment, lifestyle, and other factors. Yet, the local PRS model was less generally successful than global PRS models (with some exceptions, e.g., in Hispanic/Latino individuals), likely due to overfitting to the training dataset. In future work we will assess such models in studies with larger sample sizes. Including, it will be useful to study different potential constructions of local PRSs. While we here used LD regions, PRSs based on specific pathways, which have been recently studied [29–31], can also be viewed as local PRSs. Thus, both the definition of “local” and the method to construct local PRSs should be assessed.

It is important to highlight the differences in results when considering genetic (residual prediction, after accounting for non-genetic covariates) versus ensemble (BP phenotype prediction) model performance: the highest PVE on the phenotypic level was achieved in Black individuals. On the other hand, at the genetic level, the highest PVE was reached in White individuals, and lowest in Black individuals. We interpret this as Black participants included in the TOPMed program having the most diverse distribution in some risk factors for BP, such that the variance was well explained by the baseline covariates. This is consistent with results we recently published, showing that BP PRS performance in All of Us varied by strata of BP risk factors [13]. This further underscores the importance of incorporating multiple risk factors when developing genetic BP prediction models.

Calibration of prediction models to new populations is a well-recognized problem [32, 33]. For example, in the context of atherosclerotic cardiovascular disease, many publications suggested re-calibration of the pooled cohort equations for specific or modern populations [34, 35], including, recent literature studied the potential addition of a PRS for coronary artery disease to the pooled cohort equations, with model recalibration (for example [36, 37]). In the context of ML models in the medical literature, model calibration is offered to address model “drift”, where patient characteristics or prevalence/incidence of outcomes change over time [38].

Here, we attempted a new way to calibrate the models developed in TOPMed to the MGB Biobank dataset by a full refitting of the baseline model (only covariates), but with no update to the genetic model. Indeed, the (TOPMed-trained) genetic model performed better on the MGB Biobank dataset when the baseline model was fit on an MGB-trained baseline model. Still, more comprehensive work is needed to evaluate this and other calibration approaches in the context of genetic models, especially in the context of interactions. An important limitation of the assessment of prediction model over BP phenotypes is that BP has circadian rhythm [39], and while typically BP is measured in cohort studies early in the day, in the hospital settings it is measures at the time of the patient visit, adding noise to the MGB dataset. In our work, TOPMed-trained genetic models’ performance was lower in MGB Biobank than in TOPMed, consistent with previous results studying hypertension PRS [40]. Surprisingly, models’ performance in the subgroup of self-reported Asian individuals were high and similar to the performance in the TOPMed test dataset. However, as the number of Asian participants is low, the confidence intervals of the PVE estimates in this group are wide, limiting conclusions.

One of the strengths of this work is the use of primary, TOPMed dataset, which is comprised of large, racial/ethnically diverse and prospectively collected data used for both training and testing of the models. The dataset is of high-quality deep sequencing, joint allele-calling and the phenotypes were harmonized across studies. We used independent training and testing datasets, ensuring the reliability of model validation results. This study has some limitations as described above, including, limited sample size relative to the number of features when fitting a genetic model using local PRSs, MGB Biobank dataset is in general a “noisier” dataset, as it relies on data from health care visits and thus suffers from limitation of such datasets (BP measures may not follow best standards, may be measured using sick rather than healthy visits, etc.).

In summary, we constructed and evaluated non-linear genetic association models with SBP and DBP, composed of sequentially-trained ensembles of a baseline and a genetic model. We showed that using multiple PRSs for the same trait based on the same GWAS improves the genetic model, and further including multiple PRSs based on the same trait based on multiple GWAS further improves the model. These improvements are mostly in non-White, i.e., self-reported Black, Asian, and Hispanic, populations. We also proposed a new way to leverage ensemble dataset to calibrate a model to a new dataset: by refitting one component of the model and using the other component as it was previously trained. Our results point to the promising potential of non-linear ML to combine traditional epidemiological risk factors for hypertension with genetic score for BP prediction

## Methods

We used two datasets. The primary dataset is from the Trans-Omics in Precision Medicine (TOPMed) consortium, which was used to train and evaluate multiple models. The second dataset is from the Mass General Brigham (MGB) Biobank and was used to evaluate selected model performance in an independent healthcare-system dataset. Below we describe the datasets and the steps for constructing and evaluating models. Figure 1 describes the framework for the development of multi-PRS ML model allowing for non-genetic components to be calibrated across datasets.

### The TOPMed dataset

The TOPMed study population included 62,295 unrelated (3rd degree or less) participants from 15 studies based in the U.S. and in Taiwan. Data was extracted from the freeze 8 TOPMed dataset release. Information about the studies including ethics statements is provided in Supplementary Note 1. Blood pressure phenotypes were harmonized by the TOPMed Data Coordinating Center (DCC) [41] and included systolic blood pressure (SBP), diastolic blood pressure (DBP), and status of antihypertensive medication use (HTNMED_V1). Medication status was used to increase values of SBP and DBP by 15 and 10mmHg, respectively, to account for the expectations that their values would be higher if the corresponding individuals did not use antihypertensive medications.

### Genotype data

We used whole genome sequencing (WGS) data from Trans-Omics for Prevision Medicine (TOPMed) program [42] Freeze 8 dataset. Genome samples were sequenced through TOPMed and the National Human Genome Research Institute’s Centers for Common Disease Genomics (CCDG) program and harmonized together via joint allele calling. The methods for TOPMed WGS data acquisition and quality control (QC) are described in https://www.nhlbiwgs.org/topmed-whole-genome-sequencing-methods-freeze-8. Genetic Principal Components (PCs) and kinship coefficients were computed for the genetic data by the TOPMed DCC using the PC-Relate and PR-AiR algorithms [43] [44] implemented in the GENESIS R package [45]. Based on the kinship coefficients, we identified related individuals and generated a dataset in which all individuals were degree-3 unrelated, i.e., all kinship coefficients were <2^(−9/2)^ (approximately 0.04).We extracted allele counts of variants that passed TOPMed quality control flags from GDS files using the SeqArray package version 1.28.1 and then further processed the genetic data using custom R version 3.6.2 and Python version 3.6.15 scripts. Only variants with minor allele ≥ 0.01 in the TOPMed dataset and that passed TOPMed QC were used in this study.

### Blood pressure phenotypes

We trained ML models to predict two phenotypes: systolic blood pressure (SBP) and diastolic blood pressure (DBP). SBP and DBP values were extracted, when available, from a harmonized datasets created by TOPMed [41], and for some TOPMed-parent studies they were prepared by study researchers. We removed individuals with outlying values, defined by phenotypic values above the 99^th^ quantile and values below the 1^st^ quantile for each of the phenotypes, computed over the complete dataset. Values for SBP and DBP for individuals that were taking antihypertensive medications were adjusted by increasing SBP and DBP values by 15 and 10mmHg, respectively.

### Summary statistics from published GWAS

We used summary statistics from published GWAS of SBP and DBP provided by BioBank Japan Project (BBJ), Million Veteran Program (MVP), as well as UK BioBank and the International Consortium of Blood-Pressure Genome Wide Associations Studies (UKBB + ICBP). Details are provided in Supplementary Table 10. Because some of TOPMed European ancestry participants also participated in the UKBB+ICBP meta-analysis, we performed GWAS of SBP and DBP using all self-reported White individuals in the TOPMed dataset, and then applied a procedure to “remove” the contribution of this GWAS from the overall UKBB+ICBP GWAS summary statistics [46] as described in Supplementary Note 2.

### Polygenic Risk Scores

We developed two types of PRS – “global PRS”, using SNPs from the entire genome, and, in secondary analysis, “local PRS”, calculated from SNPs within LD-regions. In both cases SNPs were selected using the clump & threshold approach. We developed and constructed global PRSs using PRSice2 version 2.3.5 [47], from the BP GWAS summary statistics described above. As tuning parameters, we set R^2^ = 0.1, distance = 1000 kB, and several p-value thresholds: 5×10^-8^, 10^-7^, 10^-6^, …, 10^-2^. We used the TOPMed data set as a reference panel for LD (used for clumping). In secondary analysis, we developed local PRSs. We used previously computed LD-regions [48] and provided in BED files defining chromosomal segments (see Data Availability) based on a European reference panel to subset the UKB+ICBP GWAS summary statistics to files consisting of variants summary statistics for variants falling within each LD-region (chromosomal segment). We developed local PRS based for each of these regions using the same clump & threshold approach using PRSice2, but now, due to the large number of segments and thus features, we used a single p-value threshold of 10^-2^. For this secondary analysis we only used the UKB+ICBP GWAS because we saw before that, although it is based on single genetic ancestry (European) PRS based on it perform well when evaluated in various self-reported race/ethnic groups [13]. In a Supplementary Note 3 we also describe the comparison of the models’ performance using global SBP and DBP developed using PRS-CSx [49], which is an extension of the Bayesian polygenic prediction method PRS-CS [50], from the three GWAS summary statistics used in this study.

### Non-linear ML model training and hyperparameter tuning

We used the python version 3.6.15 library xgboost version 1.5.2 to fit ensembles of gradient boosted trees. We performed hyperparameter tuning using Optuna [51] library version 3.0.6. Specifically, we split the training dataset at random into 5 independent datasets and performed a 5-fold cross-validation procedure to select optimal values of relevant tuning parameters.

### Ensemble Model

As described in Figure 1, the ensemble ML model consisted of two consecutive components – “baseline model” and “genetic model”. The goal behind the two-model construction was (1) separately assess the benefit of non-linear modelling of non-genetic measures and genetic measures, (2) allow for flexible combination of the two models (e.g., linear model for covariates and non-linear model for PRSs, or the other way around), and (3) facilitate model calibration and generalizability between datasets while acknowledging that some covariates are potentially dataset-specific, and these are included only in the baseline model. The genetic model is expected to be fully transferable to a new dataset by using features that are comparable (or harmonized) across datasets.

We compared two potential constructions of both the baseline and the genetic models: non-linear ML (allowing for data-driven incorporation of interactions), and linear regression (without modeling interactions). We divided the dataset such that 70% of the data was used as training data and 30% of the data was held out as a validation set. First, we trained the baseline model using covariates only, including age, sex, self-reported race/ethnicity, BMI, and study.

The genetic model was trained on the same set of features as in the baseline model, other than study, which is dataset-specific, and it also included genetic components, i.e., PRSs, where we used the global PRSs described above, the local PRSs in secondary analysis (described later). The genetic model was trained to predict the residuals from the baseline model.

### Model development using multiple PRSs

In primary analysis, we studied the use of multiple PRSs in the genetic model via multiple models of increased complexity (in the sense that they include higher number of PRSs). We refer to these models as Level 1, Level 2 and Level 3 with Level X being shorthand for Model complexity levels (Figure 1 c). Note that all genetic models included the same non-genetic covariates as described above, and they only differed by the inclusion of additional PRSs. Level 1 of the genetic model included a single PRS developed from the UKBB+ICBP GWAS summary statistics using the p-value threshold 10^-2^. Level 2 included 7 PRSs, all from the UKBB+ICBP GWAS, and based on all considered p-value threshold: 5×10^-8^, 10^-7^, 10^-6^, …, 10^-2^. Level 3 included PRSs constructed based on all considered GWAS (UKBB+ICBP, MVP, and BBJ) GWASs and using all p-value thresholds, i.e., it included 21 PRSs.

### Secondary analysis using local PRSs

We considered using local PRSs instead of global PRSs because they may result in more interpretable models, i.e., where one could hopefully explain why different genomic regions may have different potential contributions to the BP model. Due to the large number of PRSs when computed over all LD regions, potentially resulting in model overfitting, we augmented the ensemble model approach with a variable selection step. Here, we applied LASSO regression from the python library scikit-learn version 0.24.2 using, as before, cross-validation for tuning parameter selection, on a linear model predicting the residuals from the baseline model using all constructed local PRSs. We next use the selected PRSs in the genetic model (see Figure 4a for visualization of the Ensemble model with integrated LASSO step). We evaluated this model performance on the test dataset, and also, for comparison, of the model that uses only the LASSO (without the following non-linear XGBoost ML genetic model).

### Model performance evaluation

Models’ performance was assessed on the test dataset (30% of the TOPMed dataset of unrelated individuals). We report two performance measures: percent variance explained (PVE) at both the residual (of the baseline model) and at the phenotypic (original BP phenotypes) level. To explain how each is computed we introduce some notation. Including, we distinguish between the predicted values of the baseline model, the genetic model, and the combined ensemble, and between the residuals of the baseline and the genetic models.

Let *M*_b_ and *M*_g_ denote a baseline and a genetic model, *x*_*b*_ and *x*_*g*_ the sets of covariates used by *M*_b_, and *M*_g_ respectively, and *g* the set of PRSs used by *M*_g_. Let *y* denote a BP outcome. A trained model *M*_b_ uses *x*_*b*_ to predict *y*, as:

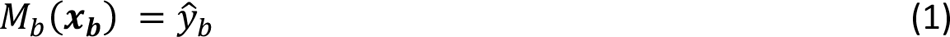

With *y*’_b_ being the prediction of *M*_b_ applied on *x*_*b*_. The residuals of model *M*_b_are obtained as the difference between the observed and the predicted BP value, and are denoted by *r*_b_:

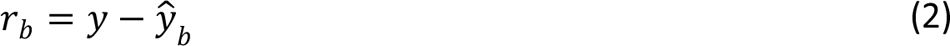

The genetic model is trained to predict *r*_b_ using *x*_*g*_ and PRSs *g*. Thus

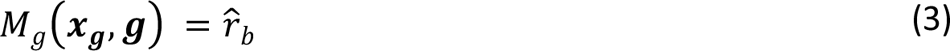

The residuals of *M*_g_ are given as the difference between the value it attempts to predict and its prediction:

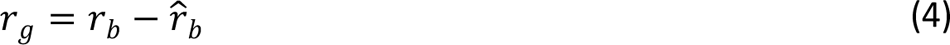

Noting based on equations (2) and (4) that the observed BP measure *y* can be decomposed as:

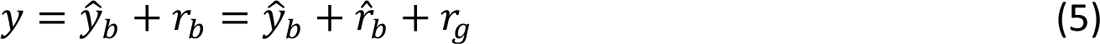

we denote the prediction BP based on the ensemble model *M*_e_ by *y*’_ensemble_ with:

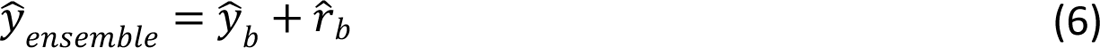

with the residuals *r*_g_of the genetic models being also the residuals of the ensemble model. For a given outcome *out* the PVE by the predicted outcome *out*^ is defined as the percent reduction in the variance of *out* when accounting for *out*^, using this formula:

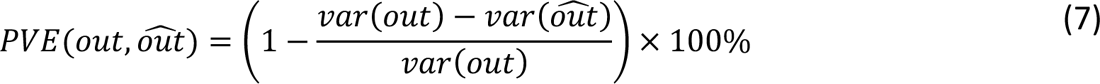

Finally, we define the performance measures of the various models. To assess the performance of baseline models we compute phenotyping PVE, *PVE*(*y*, *y*’_b_). To assess the performance of the genetic models we compute the PVE at the level of the baseline model residuals, i.e., *PVE*(*r*_b_, *r*^_b_). To assess the performance of the ensemble model we compute again PVE at the phenotypic level, i.e., *PVE*(*y*, *y*’_ensemble_).

To further interpret results, we computed 95% confidence intervals for the estimated PVEs using bootstrap sampling from the test dataset, with 100 repetitions. Specifically, we sampled with replacement individuals from the test dataset using the same sample size of the test dataset relevant for each analysis, and applied the prediction models that were trained over the train dataset. When assessing genetic model, we applied both the baseline and the genetic model over the sampled individuals. The PVE was computed for each bootstrap sample, and the 95% confidence interval was derived using the percentile method, i.e. using the 2.5 and 97.5 percentiles of the bootstrap distributions.

### The Mass General Brigham (MGB) Biobank dataset

To assess how the patterns observed in the TOPMed dataset generalize to a healthcare-based medical system, we implemented all fitted models on the MGB Biobank dataset (MGB Biobank). In MGB Biobank, phenotypes are available from electronic health records (EHR). Because blood pressure, BMI, and medication data are available sporadically depending on patient visits, we restricted the datasets health records from two years, 5/25/2021 to 5/25/2023, so that all time-varying variable correspond to approximately the same age and time. The initial MGB Biobank dataset included data for 142,476 individuals. Of these, N = 108,389 individuals did not take antihypertensive medications (dataset codes “antihypertensive medications”, “antihypertensive-other”, “beta blockers/related”, “calcium channel blockers”, “diuretics”, “direct renin inhibitor”, “antihypertensive combinations”). Next, we further filtered the dataset to only include individuals with available systolic or diastolic reading (N = 29,282). We then only included individuals with genetic data, who also are genetically unrelated to each other, resulting in 9,494 individuals in the final dataset. For each individual, we extracted the median SBP, DBP, and BMI values from these years. Individuals self-identified with categories of race and ethnicity. Individuals with Hispanic ethnicity were set to the “Hispanic” category, and otherwise individuals with non-Hispanic ethnicity and with Black or African American race to Black, and those with non-Hispanic White or Asian race were set to White or Asian, respectively, and non-Hispanic individuals with more than one self-identified race or with “other” or “unknown” were set to “other”. More details about the MGB Biobank dataset are provided in Supplementary Note 4.

To calibrate the ensemble model to the MGB Biobank dataset, we trained the baseline model on the set of covariates from MGB Biobank for prediction of the phenotype (SBP/DBP) and calculated the residuals. Next, we evaluated, separately, the genetic model (trained on the TOPMed training dataset) and the ensemble (MGB Biobank-trained baseline model + TOPMed trained genetic model) models. The genetic models included the three model complexity levels trained on the TOPMed dataset. To assess this calibration approach, we assessed the performance of the TOPMed-trained genetic model when applied over residuals from the MGB-trained and residuals from the TOPMed-trained baseline models.

## Data availability

TOPMed freeze 8 WGS data and harmonized BP phenotypes are available by application to dbGaP according to the study specific accessions: Amish: “<u>phs000956</u>”, ARIC: “<u>phs001211</u>“, BioMe: “<u>phs001644</u>”, CARDIA: “<u>phs001612</u>”, CFS: “<u>phs000954</u>”, CHS: “<u>phs001368</u>”, COPDGene: “<u>phs000951</u>”, FHS: “<u>phs000974</u>”, GENOA: “<u>phs001345</u>”, GenSalt: “<u>phs001217</u>”, HCHS/SOL: “<u>phs001395”</u>, JHS: “<u>phs000964</u>”, MESA: “<u>phs001211</u>”, THRV: “<u>phs001387</u>”, WHI: “<u>phs001237</u>”. Summary statistics from MVP BP GWAS are available from dbGaP by application to study accession “<u>phs001672</u>”. The summary statistics from the UKBB + ICBP BP GWAS are available at https://grasp.nhlbi.nih.gov/FullResults.aspx. MGB Biobank genotyping and phenotypic data are available to Mass General Brigham investigators with required approval from the Mass General Brigham Institutional Review board (IRB). Data needed to construct the selected BP PRSs generated in this study will become publicly available on a Zenodo repository upon paper acceptance and will include variants, alleles, and weights for each of the PRS based on GWAS of SBP and DBP. The BED files that define LD-regions used for construction of local PRSs are available under the Bitbucket repository in https://bitbucket.org/nygcresearch/ldetect-xsdata/src/master/.

## Acknowledgements

This study was performed as a collaboration of the NHLBI Trans-Omics in Precision Medicine (TOPMed) Consortium. We gratefully acknowledge the studies and participants who provided biological samples and data for TOPMed and CCDG. TOPMed and CCDG acknowledgements, as well as descriptions, acknowledgements, and ethics statements of contributing studies are provided in Supplementary Note 5. TOPMed consortium researchers and their affiliations are listed in Supplementary Note 6. The views expressed in this manuscript are those of the authors and do not necessarily represent the views of the National Heart, Lung, and Blood Institute; the National Institutes of Health; or the U.S. Department of Health and Human Services. We thank Mass General Brigham Biobank for providing samples, genomic data, and health information data. We also thank the HPC support team of Enterprise Research Infrastructure & Services at Mass General Brigham for their support and for the provision of computational resources. This work was approved by the Mass General Brigham Institutional Review Board and by the Beth Israel Deaconess Medical Center Committee on Clinical Investigations. This study was supported by National Heart Lung and Blood Institute (NHLBI) grant R01HL161012 to TS. MM was supported NHLBI grant K08HL159318.

## Code availability

Jupyter notebooks used for analysis in this manuscript (not including data) will be made available on GitHub upon paper acceptance.

## Author contribution statement

YH and TS drafted the manuscript. BS and ME developed PRSs and developed association models. YH and BS prepared figures and tables. NK, BS preprocessed and harmonized analytic datasets. GL advised the developed of ensemble models. TO, TK, MF, DL-J, JB, BMP, XG, WK, MM, PC, BCJ, EC, APC, SR, YDIC, CG, R-HC, CK, RC, BH, JS, SK, WZ, RJFL, BDM, RK, SSR, JIR, DL, ACM contributed to design and data curation in TOPMed studies they represent. BS, ME, NK, GL, TO, TK, MF, DL-J, JB, BMP, NS, XG, WK, MM, PC, BCJ, EC, APC, SR, YDIC, CG, R-HC, CK, RC, BH, JS, SK, WZ, RJFL, BDM, RK, SSR, JIR, DL, ACM critically reviewed the manuscript.

