## Supplemental Material for "Machine learning models for blood pressure phenotypes combining multiple polygenic risk scores"

### Supplementary Tables

#### Supplementary Table 1: Participant characteristics of the TOPMed training dataset.

| Characteristic | White | Black | Hispanic/Latino | Asian | Other/Unknown |
|---|---|---|---|---|---|
| **N** | 19,829 | 10,961 | 8,018 | 4,267 | 481 |
| **Gender**[1] | | | | | |
| Female | 12,310 (62%) | 6,713 (61%) | 4,838 (60%) | 2,143 (50%) | 198 (41%) |
| Male | 7,519 (38%) | 4,248 (39%) | 3,180 (40%) | 2,124 (50%) | 283 (59%) |
| **Age**[2] | 59 (48, 68) | 54 (47, 63) | 53 (43, 62) | 47 (39, 55) | 59 (50, 67) |
| **SBP**[2] | 125 (113, 141) | 133 (119, 150) | 127 (113, 146) | 122 (110, 137) | 134 (116, 152) |
| **DBP**[2] | 75 (68, 83) | 80 (72, 89) | 76 (68, 85) | 75 (68, 85) | 76 (66, 86) |
| **BMI**[2] | 26.5 (23.6, 30.0) | 29.1 (25.2, 34.0) | 29.0 (25.9, 33.0) | 23.9 (21.7, 26.2) | 27.3 (24.3, 31.5) |
| **Hypertensive**[1] | 11,031 (56%) | 7,722 (70%) | 4,696 (59%) | 2,160 (51%) | 323 (67%) |

**[1]n (%)**
**[2]Median (IQR)**

TOPMed training dataset characteristics combined over the studies broken into race/ethnicity background.

#### Supplementary Table 2: Participant characteristics of the TOPMed testing dataset.

| Characteristic | White | Black | Hispanic/Latino | Asian |
|---|---|---|---|---|
| **N** | 10,839 | 3,626 | 3,886 | 388 |
| **Gender**[1] | | | | |
| Female | 7,908 (73%) | 2,452 (68%) | 2,270 (58%) | 233 (60%) |
| Male | 2,931 (27%) | 1,174 (32%) | 1,616 (42%) | 155 (40%) |
| **Age**[2] | 62 (52, 70) | 57 (45, 66) | 51 (42, 60) | 63 (55, 70) |
| **SBP**[2] | 127 (113, 144) | 132 (116, 150) | 125 (113, 141) | 128 (111, 149) |
| **DBP**[2] | 75 (68, 83) | 79 (71, 88) | 76 (68, 84) | 76 (68, 84) |
| **BMI**[2] | 26.2 (23.4, 30.0) | 29.0 (25.0, 33.6) | 29.0 (26.0, 33.0) | 24.0 (22.0, 27.0) |
| **Hypertensive**[1] | 6,243 (58%) | 2,381 (66%) | 2,037 (52%) | 221 (57%) |

**[1]n (%)**
**[2]Median (IQR)**

TOPMed testing dataset characteristics combined over the studies broken into race/ethnicity background.

#### Supplementary Table 3: Number of TOPMed individuals contributed by parent studies.

| Study | White | Black | Hispanic / Latino | Asian | Other/Unknown |
|---|---|---|---|---|---|
| **N** | 30,668 | 14,587 | 11,904 | 4,655 | 481 |
| **Amish**[1] | 1,098 (3.6%) | 0 (0%) | 0 (0%) | 0 (0%) | 0 (0%) |
| **ARIC**[1] | 5,872 (19%) | 1,294 (8.9%) | 0 (0%) | 0 (0%) | 0 (0%) |
| **BioMe**[1] | 1,675 (5.5%) | 1,870 (13%) | 3,105 (26%) | 103 (2.2%) | 481 (100%) |
| **CARDIA**[1] | 1,647 (5.4%) | 1,368 (9.4%) | 0 (0%) | 0 (0%) | 0 (0%) |
| **CFS**[1] | 282 (0.9%) | 379 (2.6%) | 0 (0%) | 0 (0%) | 0 (0%) |
| **CHS**[1] | 2,686 (8.8%) | 654 (4.5%) | 31 (0.3%) | 0 (0%) | 0 (0%) |
| **COPDGene**[1] | 3,628 (12%) | 2,208 (15%) | 0 (0%) | 0 (0%) | 0 (0%) |
| **FHS**[1] | 3,084 (10%) | 0 (0%) | 11 (<0.1%) | 0 (0%) | 0 (0%) |
| **GENOA**[1] | 0 (0%) | 1,041 (7.1%) | 0 (0%) | 0 (0%) | 0 (0%) |
| **GenSalt**[1] | 0 (0%) | 0 (0%) | 0 (0%) | 1,791 (38%) | 0 (0%) |
| **HCHS/SOL**[1] | 0 (0%) | 0 (0%) | 7,455 (63%) | 0 (0%) | 0 (0%) |
| **JHS**[1] | 0 (0%) | 3,281 (2%) | 0 (0%) | 0 (0%) | 0 (0%) |
| **MESA**[1] | 1,811 (5.9%) | 1,079 (7.4%) | 1,001 (8.4%) | 582 (13%) | 0 (0%) |
| **THRV**[1] | 0 (0%) | 0 (0%) | 0 (0%) | 10986 (43%) | 0 (0%) |
| **WHI**[1] | 8,885 (29%) | 1,413 (9.7%) | 301 (2.5%) | 193 (4.1%) | 0 (0%) |

**[1]n (%)**

TOPMed testing dataset characteristics broken by study and race/ethnicity background.

#### Supplementary Table 4: Performance of the baseline model using global PRSs and TOPMed data.

| Phenotype | Group | Model | Training set (N) | Testing set (N) | Training PVE | Testing PVE |
|---|---|---|---|---|---|---|
| SBP | Overall | XGBoost | 43556 | 18748 | 31.28% | 30.05% |
| SBP | Black | XGBoost | 10914 | 3674 | 30.18% | 33.35% |
| SBP | Asian | XGBoost | 4281 | 374 | 25.04% | 19.69% |
| SBP | White | XGBoost | 19853 | 10823 | 29.40% | 28.93% |
| SBP | Hispanic/Latino | XGBoost | 8027 | 3877 | 31.64% | 27.66% |
| DBP | Overall | XGBoost | 43472 | 18768 | 15.93% | 17.35% |
| DBP | Black | XGBoost | 10889 | 3657 | 12.95% | 22.35% |
| DBP | Asian | XGBoost | 4220 | 403 | 17.30% | 11.89% |
| DBP | White | XGBoost | 19786 | 10877 | 13.18% | 13.38% |
| DBP | Hispanic/Latino | XGBoost | 8093 | 3831 | 13.26% | 17.75% |
| SBP | Overall | Linear Regression | 43556 | 18748 | 24.00% | 28.07% |
| SBP | Black | Linear Regression | 10914 | 3674 | 22.89% | 32.03% |
| SBP | Asian | Linear Regression | 4281 | 374 | 13.40% | 18.52% |
| SBP | White | Linear Regression | 19853 | 10823 | 22.97% | 27.22% |
| SBP | Hispanic/Latino | Linear Regression | 8027 | 3877 | 24.36% | 23.98% |
| DBP | Overall | Linear Regression | 43472 | 18768 | 11.80% | 14.31% |
| DBP | Black | Linear Regression | 10889 | 3657 | 9.01% | 18.77% |
| DBP | Asian | Linear Regression | 4220 | 403 | 10.17% | 11.54% |
| DBP | White | Linear Regression | 19786 | 10877 | 9.84% | 11.40% |
| DBP | Hispanic/Latino | Linear Regression | 8093 | 3831 | 8.70% | 12.10% |

#### Supplementary Table 5: Performance of the genetic and ensemble models using global PRSs and TOPMed data.

| Phenotype | Model | Group | Training set (N) | Testing set (N) | Training PVE Genetic model | Testing PVE Genetic model | Training PVE Ensemble model | Testing PVE Ensemble model | Model complexity level |
|---|---|---|---|---|---|---|---|---|---|
| **SBP** | XGBoost | Overall | 43556 | 18748 | 3.10% | 4.80% | 33.40% | 33.40% | Level 1 |
| **SBP** | XGBoost | Black | 10914 | 3674 | 0.80% | 0.70% | 30.80% | 33.80% | Level 1 |
| **SBP** | XGBoost | Asian | 4281 | 374 | 2.90% | 1.60% | 27.20% | 21.00% | Level 1 |
| **SBP** | XGBoost | White | 19853 | 10823 | 6.00% | 7.70% | 33.60% | 34.40% | Level 1 |
| **SBP** | XGBoost | Hispanic/Latino | 8027 | 3877 | 0.40% | 0.90% | 31.90% | 28.30% | Level 1 |
| **SBP** | XGBoost | Overall | 43556 | 18748 | 3.70% | 5.10% | 33.80% | 33.60% | Level 2 |
| **SBP** | XGBoost | Black | 10914 | 3674 | 1.20% | 1.10% | 31.00% | 34.10% | Level 2 |
| **SBP** | XGBoost | Asian | 4281 | 374 | 3.60% | 2.40% | 27.70% | 21.60% | Level 2 |
| **SBP** | XGBoost | White | 19853 | 10823 | 6.20% | 7.60% | 33.80% | 34.30% | Level 2 |
| **SBP** | XGBoost | Hispanic/Latino | 8027 | 3877 | 1.60% | 2.00% | 32.80% | 29.10% | Level 2 |
| **SBP** | XGBoost | Overall | 43556 | 18748 | 5.20% | 6.00% | 34.90% | 34.20% | Level 3 |
| **SBP** | XGBoost | Black | 10914 | 3674 | 2.60% | 2.60% | 32.00% | 35.10% | Level 3 |
| **SBP** | XGBoost | Asian | 4281 | 374 | 5.90% | 3.80% | 29.50% | 22.80% | Level 3 |
| **SBP** | XGBoost | White | 19853 | 10823 | 7.10% | 7.80% | 34.40% | 34.50% | Level 3 |
| **SBP** | XGBoost | Hispanic/Latino | 8027 | 3877 | 4.70% | 4.60% | 34.80% | 31.00% | Level 3 |
| **SBP** | Linear Regression | Overall | 43556 | 18748 | 2.80% | 4.40% | 33.20% | 33.10% | Level 1 |
| **SBP** | Linear Regression | Black | 10914 | 3674 | 0.50% | 0.80% | 30.60% | 33.90% | Level 1 |
| **SBP** | Linear Regression | Asian | 4281 | 374 | 2.50% | 1.40% | 26.90% | 20.80% | Level 1 |
| **SBP** | Linear Regression | White | 19853 | 10823 | 5.50% | 7.00% | 33.30% | 33.90% | Level 1 |
| **SBP** | Linear Regression | Hispanic/Latino | 8027 | 3877 | 0.20% | 0.90% | 31.80% | 28.30% | Level 1 |
| **SBP** | Linear Regression | Overall | 43556 | 18748 | 3.20% | 4.80% | 33.50% | 33.40% | Level 2 |
| **SBP** | Linear Regression | Black | 10914 | 3674 | 0.80% | 1.20% | 30.70% | 34.20% | Level 2 |
| **SBP** | Linear Regression | Asian | 4281 | 374 | 3.40% | 2.00% | 27.60% | 21.30% | Level 2 |
| **SBP** | Linear Regression | White | 19853 | 10823 | 5.60% | 7.10% | 33.30% | 34.00% | Level 2 |
| **SBP** | Linear Regression | Hispanic/Latino | 8027 | 3877 | 1.30% | 1.80% | 32.50% | 28.90% | Level 2 |

| | | | | | | | | | |
|---|---|---|---|---|---|---|---|---|---|
| **SBP** | Linear Regression | Overall | 43556 | 18748 | 4.30% | 6.40% | 34.20% | 34.50% | Level 3 |
| **SBP** | Linear Regression | Black | 10914 | 3674 | 1.70% | 3.20% | 31.30% | 35.50% | Level 3 |
| **SBP** | Linear Regression | Asian | 4281 | 374 | 4.90% | 3.80% | 28.70% | 22.70% | Level 3 |
| **SBP** | Linear Regression | White | 19853 | 10823 | 5.90% | 7.90% | 33.60% | 34.50% | Level 3 |
| **SBP** | Linear Regression | Hispanic/Latino | 8027 | 3877 | 4.20% | 5.70% | 34.50% | 31.70% | Level 3 |
| **DBP** | XGBoost | Overall | 43472 | 18768 | 4.20% | 4.70% | 19.40% | 21.20% | Level 1 |
| **DBP** | XGBoost | Black | 10889 | 3657 | 1.90% | 0.80% | 14.60% | 23.00% | Level 1 |
| **DBP** | XGBoost | Asian | 4220 | 403 | 5.00% | 0.90% | 21.40% | 12.70% | Level 1 |
| **DBP** | XGBoost | White | 19786 | 10877 | 6.90% | 7.30% | 19.10% | 19.70% | Level 1 |
| **DBP** | XGBoost | Hispanic/Latino | 8093 | 3831 | 1.40% | 1.50% | 14.50% | 19.00% | Level 1 |
| **DBP** | XGBoost | Overall | 43472 | 18768 | 4.50% | 4.60% | 19.70% | 21.10% | Level 2 |
| **DBP** | XGBoost | Black | 10889 | 3657 | 2.10% | 0.30% | 14.70% | 22.60% | Level 2 |
| **DBP** | XGBoost | Asian | 4220 | 403 | 5.60% | 2.90% | 22.00% | 14.40% | Level 2 |
| **DBP** | XGBoost | White | 19786 | 10877 | 6.90% | 6.70% | 19.20% | 19.20% | Level 2 |
| **DBP** | XGBoost | Hispanic/Latino | 8093 | 3831 | 2.50% | 2.70% | 15.40% | 20.00% | Level 2 |
| **DBP** | XGBoost | Overall | 43472 | 18768 | 6.50% | 5.60% | 21.40% | 22.00% | Level 3 |
| **DBP** | XGBoost | Black | 10889 | 3657 | 4.60% | 1.70% | 16.90% | 23.60% | Level 3 |
| **DBP** | XGBoost | Asian | 4220 | 403 | 7.90% | 3.40% | 23.90% | 14.90% | Level 3 |
| **DBP** | XGBoost | White | 19786 | 10877 | 8.00% | 7.20% | 20.10% | 19.60% | Level 3 |
| **DBP** | XGBoost | Hispanic/Latino | 8093 | 3831 | 5.70% | 5.00% | 18.20% | 21.90% | Level 3 |
| **DBP** | Linear Regression | Overall | 43472 | 18768 | 2.60% | 4.40% | 18.10% | 21.00% | Level 1 |
| **DBP** | Linear Regression | Black | 10889 | 3657 | 0.40% | 0.90% | 13.30% | 23.10% | Level 1 |
| **DBP** | Linear Regression | Asian | 4220 | 403 | 2.50% | 1.80% | 19.40% | 13.50% | Level 1 |
| **DBP** | Linear Regression | White | 19786 | 10877 | 5.70% | 6.80% | 18.10% | 19.30% | Level 1 |
| **DBP** | Linear Regression | Hispanic/Latino | 8093 | 3831 | -0.60% | 1.10% | 12.80% | 18.70% | Level 1 |
| **DBP** | Linear Regression | Overall | 43472 | 18768 | 2.90% | 4.50% | 18.30% | 21.10% | Level 2 |
| **DBP** | Linear Regression | Black | 10889 | 3657 | 0.90% | 1.20% | 13.70% | 23.30% | Level 2 |
| **DBP** | Linear Regression | Asian | 4220 | 403 | 3.00% | 2.70% | 19.70% | 14.30% | Level 2 |

| | | | | | | | | | |
|---|---|---|---|---|---|---|---|---|---|
| **DBP** | Linear Regression | White | 19786 | 10877 | 5.40% | 6.50% | 17.80% | 19.00% | Level 2 |
| **DBP** | Linear Regression | Hispanic/Latino | 8093 | 3831 | 0.60% | 2.40% | 13.80% | 19.70% | Level 2 |
| **DBP** | Linear Regression | Overall | 43472 | 18768 | 3.70% | 5.20% | 19.00% | 21.60% | Level 3 |
| **DBP** | Linear Regression | Black | 10889 | 3657 | 2.00% | 2.20% | 14.70% | 24.00% | Level 3 |
| **DBP** | Linear Regression | Asian | 4220 | 403 | 3.80% | 3.40% | 20.40% | 14.90% | Level 3 |
| **DBP** | Linear Regression | White | 19786 | 10877 | 5.40% | 6.60% | 17.90% | 19.10% | Level 3 |
| **DBP** | Linear Regression | Hispanic/Latino | 8093 | 3831 | 2.60% | 4.30% | 15.50% | 21.30% | Level 3 |

Performance results (attained PVEs) from the genetic models (PVEs of predicting residuals from the baseline model) and ensemble models (PVEs of predicting the raw trait) estimated in cross validation on the training dataset, and from the independent test dataset using global PRS in TOPMed dataset.

#### Supplementary Table 6: Performance of the genetic and ensemble models using global PRSs in TOPMed data developed with PRS-CSx.

| Phenotype | Model | Group | Training set (N) | Testing set (N) | Training PVE Genetic model | Testing PVE Genetic model | Training PVE Ensemble model | Testing PVE Ensemble model |
|---|---|---|---|---|---|---|---|---|
| **SBP** | Linear Regression | Overall | 43552 | 18743 | 2.11% | 3.20% | 32.75% | 32.30% |
| **SBP** | Linear Regression | Black | 10913 | 3674 | 0.54% | 1.60% | 30.59% | 34.50% |
| **SBP** | Linear Regression | Asian | 4281 | 374 | 2.34% | 2.80% | 26.77% | 22.00% |
| **SBP** | Linear Regression | White | 19850 | 10818 | 3.20% | 4.00% | 31.72% | 31.80% |
| **SBP** | Linear Regression | Hispanic/Latino | 8027 | 3877 | 1.84% | 2.30% | 32.88% | 29.30% |
| **SBP** | XGBoost | Overall | 43552 | 18743 | 2.39% | 3.20% | 32.94% | 32.30% |
| **SBP** | XGBoost | Black | 10913 | 3674 | 0.84% | 1.60% | 30.80% | 34.40% |
| **SBP** | XGBoost | Asian | 4281 | 374 | 2.56% | 2.80% | 26.93% | 22.00% |
| **SBP** | XGBoost | White | 19850 | 10818 | 3.48% | 4.10% | 31.92% | 31.80% |
| **SBP** | XGBoost | Hispanic/Latino | 8027 | 3877 | 2.12% | 2.20% | 33.07% | 29.20% |
| **DBP** | Linear Regression | Overall | 42897 | 18553 | 1.93% | 2.90% | 17.23% | 19.30% |
| **DBP** | Linear Regression | Black | 10737 | 3612 | 0.83% | 1.40% | 13.67% | 23.20% |
| **DBP** | Linear Regression | Asian | 4164 | 397 | 2.24% | 2.60% | 18.52% | 14.00% |
| **DBP** | Linear Regression | White | 19587 | 10745 | 2.83% | 3.60% | 15.29% | 16.10% |
| **DBP** | Linear Regression | Hispanic/Latino | 7939 | 3799 | 1.47% | 2.30% | 14.26% | 19.30% |
| **DBP** | XGBoost | Overall | 36883 | 12528 | 2.11% | 2.70% | 17.54% | 19.60% |
| **DBP** | XGBoost | Black | 9599 | 2486 | 1.12% | 1.70% | 14.47% | 23.70% |
| **DBP** | XGBoost | Asian | 3993 | 198 | 2.52% | 2.70% | 18.53% | 14.30% |
| **DBP** | XGBoost | White | 16241 | 7373 | 2.85% | 3.20% | 15.22% | 16.20% |
| **DBP** | XGBoost | Hispanic/Latino | 6580 | 2471 | 1.89% | 2.50% | 13.91% | 19.80% |

Performance results (attained PVEs) from the genetic models (PVEs of predicting residuals from the baseline model) and ensemble models (PVEs of predicting the raw trait) estimated in cross validation on the training dataset, and from the independent test dataset using global PRS in TOPMed dataset where PRS were developed using PRS-CSx.

#### Supplementary Table 7: Hyperparameter choice for models' training procedure.

| Phenotype | Model | n estimator | max depth | min child weight | subsample | colsample by tree | lambda | alpha | gamma | eta |
|---|---|---|---|---|---|---|---|---|---|---|
| **DBP** | TOPMed baseline model | 315 | 3 | 100 | 0.6 | 1 | 20 | 14 | 32 | 0.04 |
| **SBP** | TOPMed baseline model | 460 | 99 | 20 | 0.4 | 0.7 | 0 | 49 | 19 | 0.01 |
| **DBP** | TOPMed model 1 | 90 | 3 | 1 | 0.8 | 0.9 | 37 | 50 | 45 | 0.05 |
| **SBP** | TOPMed model 1 | 290 | 1 | 57 | 0.3 | 0.8 | 48 | 48 | 17 | 0.1 |
| **DBP** | TOPMed model 2 | 348 | 30 | 74 | 0.1 | 0.7 | 13 | 37 | 31 | 0.01 |
| **SBP** | TOPMed model 2 | 192 | 30 | 42 | 0.3 | 0.5 | 48 | 43 | 24 | 0.02 |
| **DBP** | TOPMed model 3 | 697 | 2 | 29 | 0.9 | 0.6 | 31 | 8 | 38 | 0.02 |
| **SBP** | TOPMed model 3 | 290 | 3 | 16 | 0.9 | 0.8 | 42 | 36 | 29 | 0.04 |
| **DBP** | MGB Biobank Baseline Model | 405 | 100 | 9 | 0.5 | 0.9 | 0 | 44 | 22 | 0.01 |
| **SBP** | MGB Biobank Baseline Model | 293 | 4 | 40 | 0.7 | 0.7 | 49 | 35 | 22 | 0.04 |

Hyperparameters and their value choices for models' training procedure after applying Optuna hyperparameter selection.

#### Supplementary Table 8: Performance of the genetic and ensemble models using local PRSs in TOPMed data.

| Phenotype | Model | Group | Training set (N) | Testing set (N) | Training PVE Genetic model | Testing PVE Genetic model | Training PVE Ensemble model | Testing PVE Ensemble model |
|---|---|---|---|---|---|---|---|---|
| **SBP** | Lasso | Overall | 43556 | 18748 | 6.85% | 4.52% | 36.31% | 33.21% |
| **SBP** | Lasso | Black | 10914 | 3674 | 3.76% | 1.55% | 33.20% | 34.39% |
| **SBP** | Lasso | Asian | 4281 | 374 | 5.54% | 1.40% | 29.48% | 20.82% |
| **SBP** | Lasso | White | 19853 | 10823 | 9.09% | 5.78% | 36.13% | 33.03% |
| **SBP** | Lasso | Hispanic/Latino | 8027 | 3877 | 7.04% | 4.24% | 36.77% | 30.72% |
| **SBP** | XGBoost | Overall | 43556 | 18748 | 59.27% | 1.61% | 72.15% | 31.18% |
| **SBP** | XGBoost | Black | 10914 | 3674 | 58.88% | 0.68% | 71.46% | 33.80% |
| **SBP** | XGBoost | Asian | 4281 | 374 | 58.83% | 0.28% | 69.26% | 19.92% |
| **SBP** | XGBoost | White | 19853 | 10823 | 59.45% | 2.12% | 71.51% | 30.43% |
| **SBP** | XGBoost | Hispanic/Latino | 8027 | 3877 | 59.68% | 1.62% | 72.57% | 28.83% |
| **SBP** | Linear Regression | Overall | 43556 | 18748 | 7.81% | 4.77% | 36.96% | 33.39% |
| **SBP** | Linear Regression | Black | 10914 | 3674 | 3.74% | 0.89% | 33.19% | 33.95% |
| **SBP** | Linear Regression | Asian | 4281 | 374 | 6.52% | 0.64% | 30.21% | 20.21% |
| **SBP** | Linear Regression | White | 19853 | 10823 | 10.72% | 6.76% | 37.27% | 33.73% |
| **SBP** | Linear Regression | Hispanic/Latino | 8027 | 3877 | 7.87% | 3.41% | 37.33% | 30.12% |
| **DBP** | Lasso | Overall | 43472 | 18768 | 2.69% | 2.04% | 18.19% | 19.04% |
| **DBP** | Lasso | Black | 10889 | 3657 | 1.37% | 0.69% | 14.14% | 22.89% |
| **DBP** | Lasso | Asian | 4220 | 403 | 1.86% | 1.53% | 18.84% | 13.24% |
| **DBP** | Lasso | White | 19786 | 10877 | 3.05% | 1.84% | 15.82% | 14.97% |
| **DBP** | Lasso | Hispanic/Latino | 8093 | 3831 | 4.06% | 3.95% | 16.77% | 21.00% |
| **DBP** | XGBoost | Overall | 43472 | 18768 | 83.77% | 1.93% | 86.35% | 18.94% |
| **DBP** | XGBoost | Black | 10889 | 3657 | 83.98% | 0.69% | 86.06% | 22.89% |
| **DBP** | XGBoost | Asian | 4220 | 403 | 83.83% | 2.00% | 86.63% | 13.66% |
| **DBP** | XGBoost | White | 19786 | 10877 | 83.34% | 1.74% | 85.54% | 14.89% |
| **DBP** | XGBoost | Hispanic/Latino | 8093 | 3831 | 84.31% | 3.79% | 86.39% | 20.86% |
| **DBP** | Linear Regression | Overall | 43472 | 18768 | 4.75% | 2.61% | 19.92% | 19.51% |
| **DBP** | Linear Regression | Black | 10889 | 3657 | 2.20% | 0.10% | 14.87% | 22.42% |
| **DBP** | Linear Regression | Asian | 4220 | 403 | 3.98% | 2.31% | 20.59% | 13.93% |
| **DBP** | Linear Regression | White | 19786 | 10877 | 5.76% | 2.39% | 18.17% | 15.45% |
| **DBP** | Linear Regression | Hispanic/Latino | 8093 | 3831 | 6.38% | 5.66% | 18.79% | 22.40% |

Performance results (attained PVEs) from the genetic models (PVEs of predicting residuals from the baseline model) and ensemble models (PVEs of predicting the raw trait) estimated in cross validation on the training dataset, and from the independent test dataset using local PRS in TOPMed dataset.

#### Supplementary Table 9: Participant characteristics of the MGB Biobank dataset.

| Characteristic | | White | Black | Asian | Other/Unknown |
|---|---|---|---|---|---|
| **N** | | 7,985 | 412 | 200 | 897 |
| **Gender[1]** | | | | | |
| | **Female** | 4,975 (62%) | 278 (67%) | 133 (67%) | 628 (70%) |
| | **Male** | 3,010 (38%) | 134 (33%) | 67 (34%) | 269 (30%) |
| **Age[2]** | | 61 (48, 71) | 49 (38, 60) | 49 (41, 59) | 47 (37, 60) |
| **SBP[2]** | | 124 (116, 134) | 124 (115, 132) | 119 (112, 129) | 122 (114, 132) |
| **DBP[2]** | | 74 (68, 80) | 73 (68, 80) | 73 (68, 79) | 74 (69, 80) |
| **BMI[2]** | | 26.6 (23.5, 30.5) | 29.9 (25.8, 34.9) | 24.2 (21.5, 26.7) | 28.8 (25.4, 33.3) |

**[1]n (%)**
**[2]Median (IQR)**

MGB Biobank dataset characteristics broken into self-reported race/ethnicity background of the participants.

#### Supplementary Table 10: GWAS summary statistics used for SBP and DBP PRS development.

| GWAS name | Reference | Trait | Sample size | Population |
|---|---|---|---|---|
| **BBJ** | PMID:29403010(7) | SBP | 136,597 | Japanese |
| | | DBP | 136,615 | |
| **UKBB+ICBP** | PMID:30224653 (8) | SBP | 757,601 | European |
| | | DBP | 757,601 | |
| **MVP** | PMID:30578418 (10) | SBP | 318,492 | Multi-ethnic (69.1% non-Hispanic White, 18.8% non-Hispanic Black, 6.7% Hispanic, 0.77% non-Hispanic Asian and 0.85% non-Hispanic Native American individuals) |
| | | DBP | 318,891 | |

The table provides GWAS source, study population as reported by the manuscript or repository reporting the GWAS, and number of participants used to generate summary statistics. Because some of UKBB+ICBP individuals overlapped with our TOPMed dataset, we performed GWAS using the overlapping TOPMed individuals and applied a numerical procedure (described in Supplementary Note 2 to remove their contribution from the UKBB+ICBP summary statistics.

### Supplementary Figures

#### Supplementary Figure 1: Comparison of baseline cross-validated PVE with and without inclusion of genetic PCs.

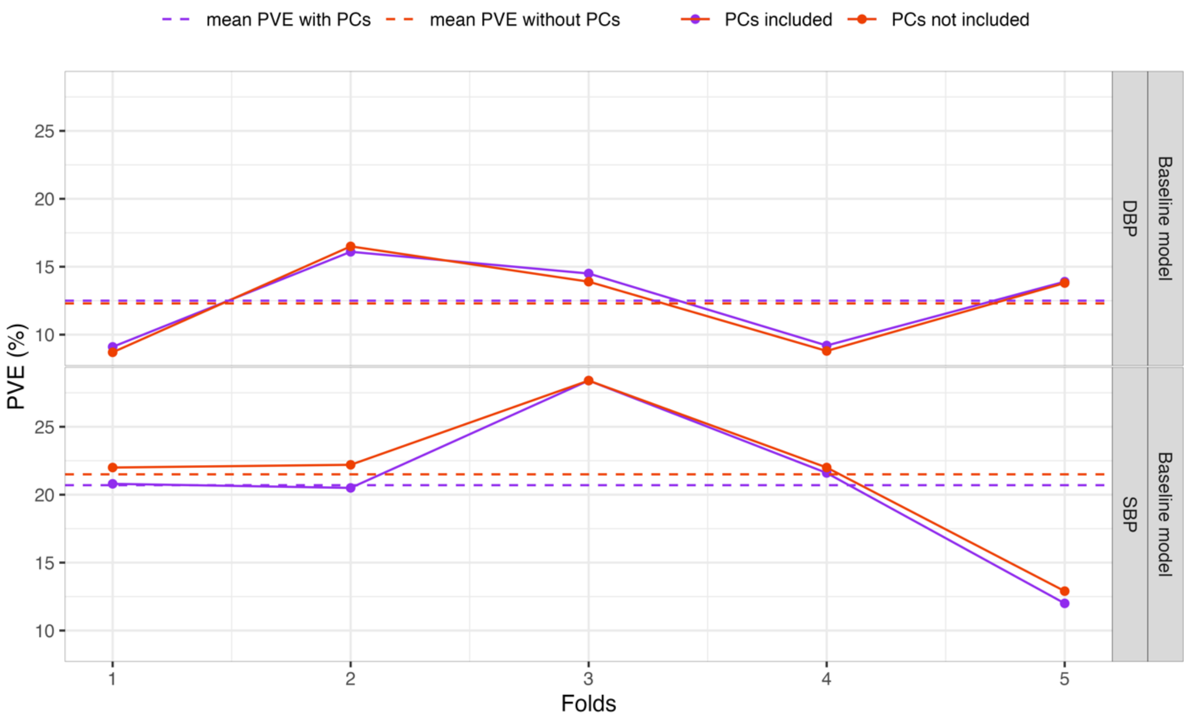

TOPMed baseline model performance for SBP and DBP prediction, comparing inclusion of genetic PCs to the model performance trained without genetic PCs.
PVE: Percent variance explained. TOPMed: Trans-Omics in Precision Medicine project. SBP: systolic blood pressure. DBP: diastolic blood pressure.

Supplementary Figure 2: Estimated PVE of genetic models fitted using XGBoost and linear models using global PRS.

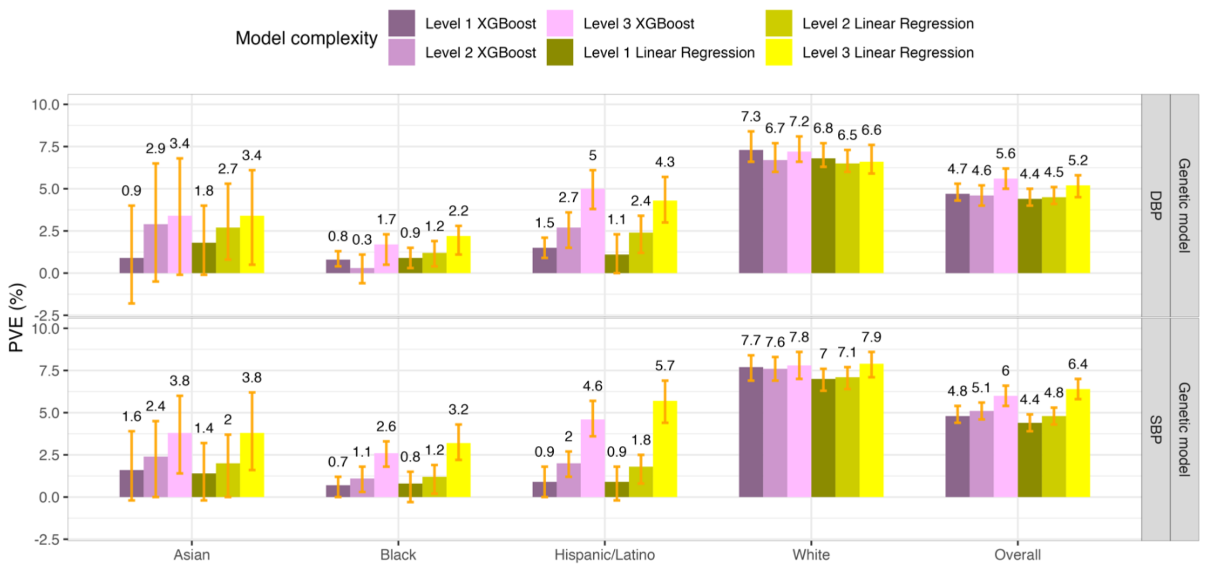

Estimated PVEs in the TOPMed test dataset for genetic model performance for prediction of SBP and DBP in the overall test dataset and stratified by self-reported race/ethnicity (White N = 10,877, Hispanic/Latino N = 3,831, Black N = 3,657, Asian N = 403 for DBP; White N = 10,823, Hispanic/Latino N = 3,877, Black N = 3,674, Asian N = 374 for SBP).

PVE: Percent variance explained. TOPMed: Trans-Omics in Precision Medicine project. SBP: systolic blood pressure. DBP: diastolic blood pressure.

Supplementary Figure 3: Performance of genetic and ensemble models fitted using XGBoost, linear regression and LASSO using local PRSs in TOPMed test dataset.

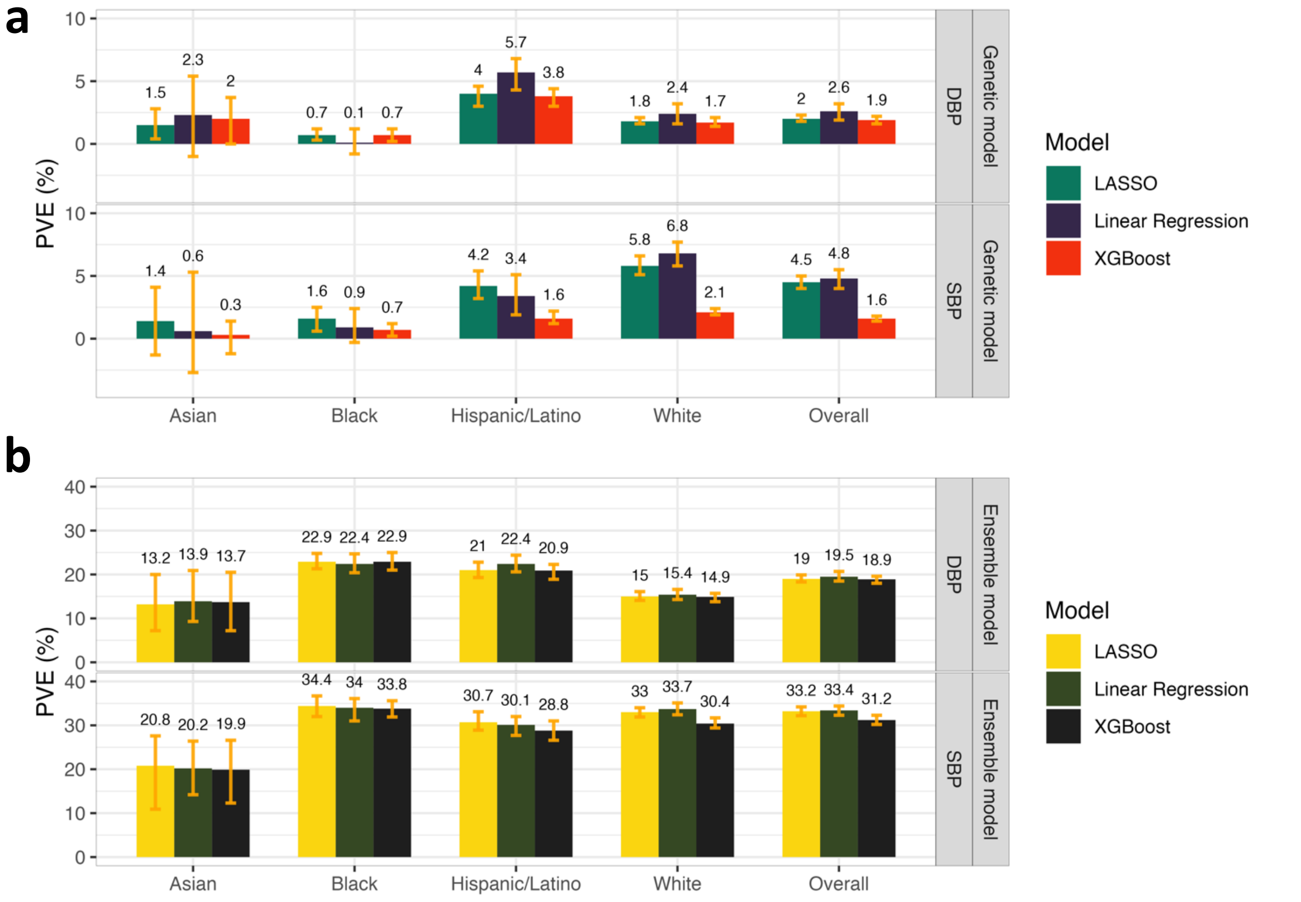

Panel a: Estimated PVEs in the TOPMed test dataset by genetic models incorporating local PRSs based on the UKB+ICBP GWAS. PVE is reported for predicting residuals from the baseline model, where the baseline model was XGBoost and only used non-genetic covariates. Panel b: Estimated PVE in the TOPMed test dataset for ensemble model at the raw phenotypic level. PVEs are reported for models of SBP and DBP, in the overall test dataset and stratified by self-reported race/ethnicity (White N = 10,877, Hispanic/Latino N = 3,831, Black N = 3,657, Asian N = 403 for DBP; White N = 10,823, Hispanic/Latino N = 3,877, Black N = 3,674, Asian N = 374 for SBP).

PVE: Percent variance explained. TOPMed: Trans-Omics in Precision Medicine project. SBP: systolic blood pressure. DBP: diastolic blood pressure. PRS: polygenic risk score.

Supplementary Figure 4: Estimated phenotypic PVE of baseline models in the MGB Biobank data fitted using XGBoost.

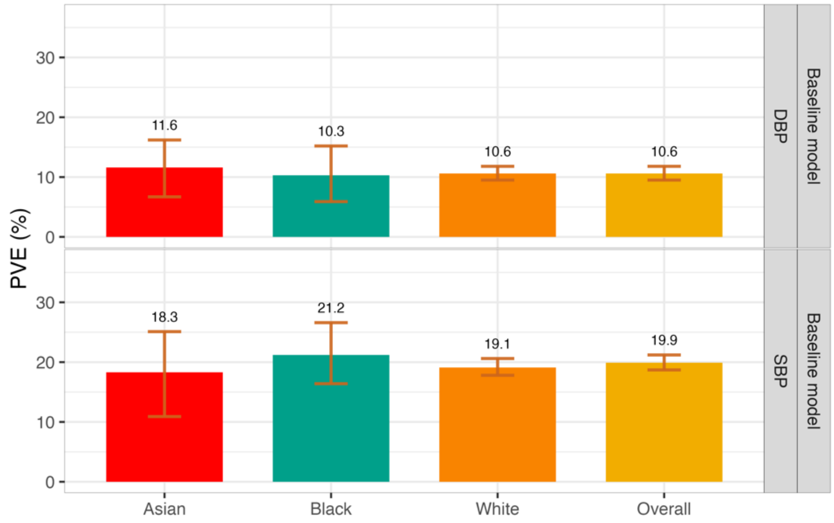

Estimated PVE in the MGBB test dataset for Baseline models' performance for prediction of SBP and DBP phenotypes in the overall test dataset and stratified by race/ethnicity (White N = 7,985, Black N = 412, Asian N = 200).

PVE: Percent variance explained. TOPMed: Trans-Omics in Precision Medicine project. SBP: systolic blood pressure. DBP: diastolic blood pressure. MGBB: Mass General Brigham Biobank.

Supplementary Figure 5: Estimated PVE of genetic and ensemble models in the MGB Biobank.

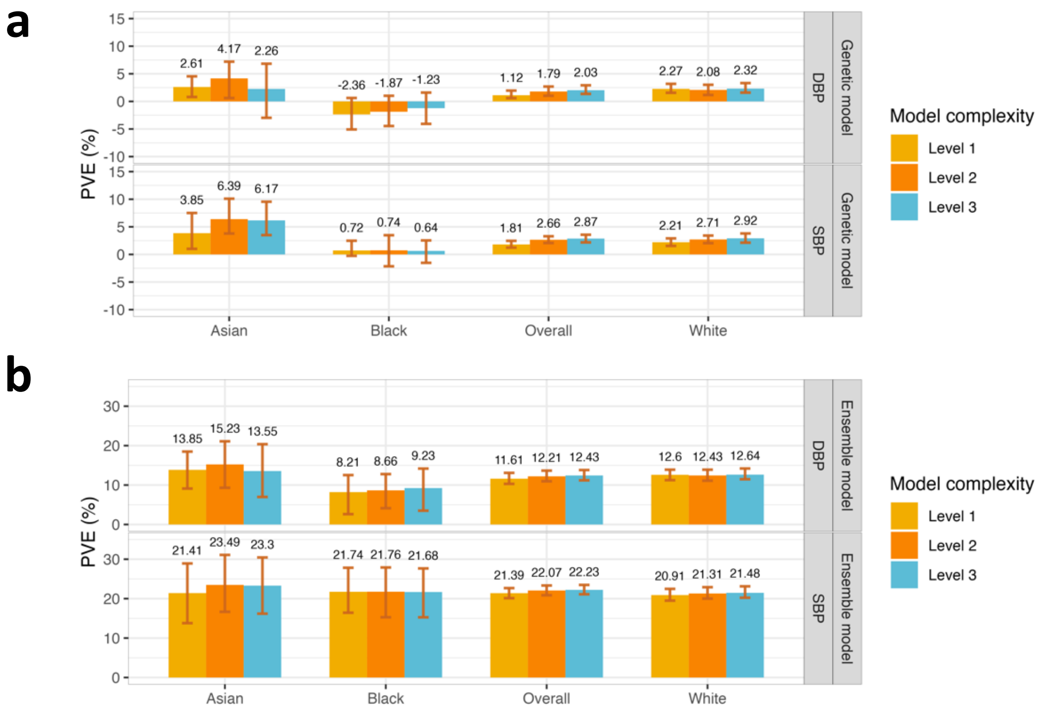

Panel a: Estimated PVE in the MGBB test dataset (including only individuals who do not take antihypertensive medication) for XGBoost Genetic models fitted on the TOPMed dataset of three levels of complexity. PVEs are shown for the performance in prediction of the second order of residuals for SBP and DBP phenotypes in the overall test dataset and stratified by race/ethnicity (White N = 7,985, Black N = 412, Asian N = 200). Panel b: Estimated PVE in the MGBB test dataset for Ensemble models' three levels of complexity performance for prediction of the SBP and DBP in the overall test dataset and stratified by race/ethnicity (White N = 7,985, Black N = 412, Asian N = 200).

PVE: Percent variance explained. TOPMed: Trans-Omics in Precision Medicine project. SBP: systolic blood pressure. DBP: diastolic blood pressure. MGBB: Mass General Brigham Biobank.

#### Supplementary Note 1: Descriptions of participating TOPMed studies

Participating TOPMed studies included: Genetics of Cardiometabolic Health in the Amish (Amish; n = 1098), Atherosclerosis Risk in Communities study (ARIC; n = 7166), Mount Sinai BioMe Biobank (BioMe; n = 7234), Coronary Artery Risk Development in Young Adults Study (CARDIA; n = 3015), Cleveland Family Study (CFS; n = 661), Cardiovascular Health Study (CHS; n = 3371), Genetic Epidemiology of COPD (COPDGene; n = 5836), Framingham Heart Study (FHS; n = 3095), Genetic Epidemiology Network of Arteriopathy (GENOA; n = 1041), Genetic Epidemiology Network of Salt Sensitivity (GenSalt, n = 1791), Hispanic Community Health Study/Study of Latinos (HCHS/SOL; n = 7455), Jackson Heart Study (JHS; n = 3281), Multi-Ethnic Study of Atherosclerosis (MESA; n= 4473), Taiwan Study of Hypertension using Rare Variants (THRV; n = 1986) and Women's Health Initiative (WHI; n = 10792). Study specific descriptions are provided below.

##### Amish

**Ethics statement:**

All study protocols were approved by the institutional review board at the University of Maryland Baltimore. Informed consent was obtained from each study participant.

**Amish acknowledgements:**

We gratefully acknowledge our Amish liaisons, research volunteers, field workers, and Amish Research Clinic staff and the extraordinary cooperation and support of the Amish community without which these studies would not have been possible. The Amish studies are supported by grants and contracts from the NIH, including U01 HL072515, U01 HL84756, U01 HL137181, and P30 DK72488. The TOPMed component of the Amish Research Program was supported by NIH grants R01 HL121007, U01 HL072515, and R01 AG18728.

#### ARIC

The Atherosclerosis Risk in Communities study (dbGaP accession phs000090) is a population-based prospective cohort study of cardiovascular disease sponsored by the National Heart, Lung, and Blood Institute (NHLBI). ARIC included 15,792 individuals, predominantly European American and African American, aged 45-64 years at baseline (1987-89), chosen by probability sampling from four US communities. Cohort members completed three additional triennial follow-up examinations, a fifth exam in 2011-2013, a sixth exam in 2016-2017, and a seventh exam in 2018-2019, and an eighth exam in 2020. The ARIC study has been described in detail previously [1, 2].

**Ethics statement:**

The ARIC study has been approved by a single Institutional Review Board (sIRB) at Johns Hopkins School of Medicine and Institutional Review Boards (IRB) at all participating institutions: University of North Carolina at Chapel Hill IRB, Johns Hopkins University School of Public Health IRB, University of Minnesota IRB, Wake Forest University Health Sciences IRB, and University of Mississippi Medical Center IRB. Study participants provided written informed consent at all study visits.

**ARIC acknowledgements:**

The Atherosclerosis Risk in Communities study has been funded in whole or in part with Federal funds from the National Heart, Lung, and Blood Institute, National Institutes of Health,

Department of Health and Human Services (contract numbers 75N92022D00001, 75N92022D00002, 75N92022D00003, 75N92022D00004, 75N92022D00005). The authors thank the staff and participants of the ARIC study for their important contributions.

WGS for “NHLBI TOPMed: Atherosclerosis Risk in Communities (ARIC)” (phs001211) was performed at the Baylor College of Medicine Human Genome Sequencing Center (HHSN268201500015C and 3U54HG003273-12S2) and the Broad Institute for MIT and Harvard (3R01HL092577- 06S1). The Genome Sequencing Program (GSP) was funded by the National Human Genome Research Institute (NHGRI), the National Heart, Lung, and Blood Institute (NHLBI), and the National Eye Institute (NEI). The GSP Coordinating Center (U24 HG008956) contributed to cross program scientific initiatives and provided logistical and general study coordination. The Centers for Common Disease Genomics (CCDG) program was supported by NHGRI and NHLBI, and whole genome sequencing was performed at the Baylor College of Medicine Human Genome Sequencing Center (UM1 HG008898).

#### BioMe

The BioMe Biobank is an ongoing, prospective, hospital- and outpatient- based population research program operated by The Charles Bronfman Institute for Personalized Medicine (IPM) at Mount Sinai. BioMe has enrolled over 50,000 participants between September 2007 and July 2019. BioMe is an Electronic Medical Record (EMR)-linked biobank that integrates research data and clinical care information for consented patients at The Mount Sinai Medical Center, which serves diverse local communities of upper Manhattan with broad health disparities. IPM BioMe populations include 25% of African American ancestry (AA), 36% of Hispanic Latino ancestry (HL), 30% of white European ancestry (EA), and 9% of other ancestry. The BioMe disease burden is reflective of health disparities in the local communities. BioMe operations are fully integrated in clinical care processes, including direct recruitment from clinical sites waiting areas and phlebotomy stations by dedicated BioMe recruiters independent of clinical care providers, prior to or following a clinician standard of care visit. Recruitment currently occurs at a broad spectrum of over 30 clinical care sites.

**Ethics statement:**

The BioMe cohort was approved by the Institutional Review Board at the Icahn School of Medicine at Mount Sinai. All BioMe participants provided written, informed consent for genomic data sharing.

**BioMe acknowledgements:**

The Mount Sinai BioMe Biobank has been supported by The Andrea and Charles Bronfman Philanthropies and in part by Federal funds from the NHLBI and NHGRI (U01HG00638001; U01HG007417; X01HL134588). We thank all participants in the Mount Sinai Biobank. We also thank all our recruiters who have assisted and continue to assist in data collection and management and are grateful for the computational resources and staff expertise provided by Scientific Computing at the Icahn School of Medicine at Mount Sinai.

#### CARDIA

The Coronary Artery Risk Development in Young Adults study (dbGaP accession phs000285) is a prospective multicenter study with 5,115 adults Caucasian and African American participants of the age group 18-30 years at baseline, recruited from four centers at the baseline examination in 1985-1986 [3]. The recruitment was done from the total community in Birmingham, AL, from selected census tracts in Chicago, IL and Minneapolis, MN; and from the Kaiser Permanente health plan membership in Oakland, CA. Nine examinations have been completed in the years 0, 2, 5, 7, 10, 15, 20, 25 and 30, with high retention rates (91%, 86%, 81%, 79%, 74%, 72%, 72%, and 71%, respectively) and written informed consent was obtained in each visit.

**Ethics statement:**

All CARDIA participants provided informed consent, and the study was approved by the Institutional Review Boards of the University of Alabama at Birmingham and the University of Texas Health Science Center at Houston.

**CARDIA acknowledgements:**

The Coronary Artery Risk Development in Young Adults Study (CARDIA) is conducted and supported by the National Heart, Lung, and Blood Institute (NHLBI) in collaboration with the University of Alabama at Birmingham (HHSN268201800005I & HHSN268201800007I), Northwestern University (HHSN268201800003I), University of Minnesota (HHSN268201800006I), and Kaiser Foundation Research Institute (HHSN268201800004I). CARDIA was also partially supported by the Intramural Research Program of the National Institute on Aging (NIA) and an intra-agency agreement between NIA and NHLBI (AG0005).

#### CFS

The Cleveland Family Study (CFS) was designed to examine the genetic basis of sleep apnea in 2,534 African-American and European-American individuals from 356 families. Index probands with confirmed sleep apnea were recruited from sleep centers in northern Ohio, supplemented with additional family members and neighborhood control families [4]. Four visits occurred between 1990 and 2006; in the first 3, data were collected in participants' homes while the last occurred in a clinical research center (2000 - 2006). Measurements included sleep apnea monitoring, blood pressure, anthropometry, spirometry and other related phenotypes. Blood samples (overnight fasting, before bed and following an oral glucose tolerance test), nasal and oral ultrasound, and ECG were also obtained during the 4th exam. Institutional Review Board approval and signed informed consent was obtained for all participants.

**Ethics statement:**

Cleveland Family Study was approved by the Institutional Review Board (IRB) of Case Western Reserve University and Mass General Brigham (formerly Partners HealthCare). Written informed consent was obtained from all participants.

#### CHS

The Cardiovascular Health Study (CHS) is a population-based cohort study initiated by the National Heart, Lung and Blood Institute (NHLBI) in 1987 to determine the risk factors for development and progression of cardiovascular disease (CVD) in older adults, with an emphasis on subclinical measures. The study recruited 5,888 adults aged 65 or older at entry in four U.S. communities and conducted extensive annual clinical exams between 1989-1999 along with semi-annual phone calls, events adjudication, and subsequent data analyses and publications. Additional data are collected by studies ancillary to CHS. In June 1990, four Field Centers (Sacramento, CA; Hagerstown, MD; Winston-Salem, NC; Pittsburgh, PA) completed the recruitment of 5201 participants. Between November 1992 and June 1993, an additional 687 adults of primarily African Americans ethnicity were recruited using similar methods.

Blood samples were drawn from all participants at their baseline examination and during follow-up clinic visits and DNA was subsequently extracted from available samples. CHS analyses were limited to participants with available DNA who consented to genetic studies. The baseline examinations consisted of a home interview and a clinic examination that assessed not only traditional risk factors but also measures of subclinical disease, including carotid ultrasound, echocardiography, electrocardiography, and pulmonary function. Between enrollment and 1998-99, participants were seen in the clinic annually, and contacted by phone at 6-month intervals to collect information about hospitalizations and potential cardiovascular events. Major exam components were repeated during annual follow-up examinations through 1999. Cranial MRI scans, retinal photography, and tests of endothelial function were added as new components. Standard protocols for the identification and adjudication of events were implemented during follow-up. The adjudicated events are CHD, angina, heart failure (HF), stroke, transient ischemic attack (TIA), claudication and mortality. Adjudication of cause of death continues using a streamlined protocol; adjudication of other events ended in June 2015. Deep venous thrombosis and pulmonary embolism events from baseline through 2001 were adjudicated in an ancillary study: the Longitudinal Investigation of Thromboembolism Etiology (LITE). Since 1999, participants have been contacted every 6 months by phone, primarily to ascertain health status and for events follow-up. The study was initially approved by institutional review boards at the Field Centers (Wake Forest, University of California – Davis, Johns Hopkins University, University of Pittsburgh), the Core Laboratory (University of Vermont) and at the Coordinating Center (University of Washington). The University of Washington now handles CHS Data Repository approvals.

**Ethics statement:**

All CHS participants provided informed consent, and the study was approved by the Institutional Review Board [or ethics review committee] of University Washington.

**CHS acknowledgements:**

Cardiovascular Health Study: This research was supported by contracts HHSN268201200036C, HHSN268200800007C, HHSN268201800001C, N01HC55222, N01HC85079, N01HC85080, N01HC85081, N01HC85082, N01HC85083, N01HC85086, 75N92021D00006, and grants U01HL080295, U01HL130114, and HL105756 from the National Heart, Lung, and Blood Institute

(NHLBI), with additional contribution from the National Institute of Neurological Disorders and Stroke (NINDS). Additional support was provided by R01AG023629 from the National Institute on Aging (NIA). A full list of principal CHS investigators and institutions can be found at CHS-NHLBI.org. The content is solely the responsibility of the authors and does not necessarily represent the official views of the National Institutes of Health.

#### COPDGene

COPDGene [5] is a cohort study for respiratory disease research, recruiting more than 10,000 subjects between the ages of 45 and 80 who had at least 10 pack-years of smoking during January 2008 - June 2011 at 21 clinical centers. Participants were characterized using spirometry, six-minute walk, inspiratory and expiratory chest CT scans, respiratory symptoms, medical history, medication history and 36-Item short form health survey. In the current analysis, we only used COPDGene control participants (meaning, individuals without COPD).

**Ethics statement:**

All COPDGene participants provided written informed consent, and the study was approved by the Institutional Review Boards of the participating clinical centers.

**COPDGene acknowledgements:**

The COPDGene project described was supported by Award Number U01 HL089897 and Award Number U01 HL089856 from the National Heart, Lung, and Blood Institute. The content is solely the responsibility of the authors and does not necessarily represent the official views of the National Heart, Lung, and Blood Institute or the National Institutes of Health. The COPDGene project is also supported by the COPD Foundation through contributions made to an Industry Advisory Board comprised of AstraZeneca, Boehringer Ingelheim, GlaxoSmithKline, Novartis,

Pfizer, Siemens and Sunovion. A full listing of COPDGene investigators can be found at: http://www.copdgene.org/directory

#### FHS

The Framingham Heart Study (dbGaP accession phs000007) began in 1948 with the recruitment of an original cohort of 5,209 men and women (mean age 44 years; 55 percent women). In 1971 a second generation of study participants was enrolled; this cohort (mean age 37 years; 52% women) consisted of 5,124 children and spouses of children of the original cohort. A third-generation cohort of 4,095 children of offspring cohort participants (mean age 40 years; 53 percent women) was enrolled in 2002-2005 and are seen every 4 to 8 years. Details of study designs for the three cohorts are summarized elsewhere [6-8]. At each clinic visit, a medical history was obtained, and participants underwent a physical examination. Only study participants consented for genetic and non-genetic data are included. FHS has been approved by the Boston University IRB

**Ethics statement:**

The Framingham Heart Study was approved by the Institutional Review Board of the Boston University Medical Center. All study participants provided written informed consent.

**FHS acknowledgements:**

The Framingham Heart Study (FHS) acknowledges the support of contracts NO1-HC-25195, HHSN268201500001I and 75N92019D00031 from the National Heart, Lung and Blood Institute and grant supplement R01 HL092577-06S1 for this research. We also acknowledge the dedication of the FHS study participants without whom this research would not be possible. Dr. Vasan is supported in part by the Evans Medical Foundation and the Jay and Louis Coffman Endowment from the Department of Medicine, Boston University School of Medicine.

#### GENOA

The Genetic Epidemiology Network of Arteriopathy (GENOA) study (dbGaP accession phs000379), a part of the Family Blood Pressure Program (FBPP Investigators, 2002), consists of hypertensive sibships that were recruited for linkage and association studies in order to identify genes that influence blood pressure and its target organ damage (Daniels, 2004). In the initial phase of the GENOA study (Phase I: 1996-2001), all members of sibships containing ≥ 2 individuals with essential hypertension clinically diagnosed before age 60 were invited to participate, including both hypertensive and normotensive siblings. In the second phase of the GENOA study (Phase II: 2000-2004), 1,239 non-Hispanic white and 1,482 African American participants were successfully re-recruited to measure potential target organ damage due to hypertension.

**Ethics statement:**

Written informed consent was obtained from all subjects and approval was granted by participating institutional review boards (University of Michigan, University of Mississippi Medical Center, and Mayo Clinic).

**GENOA acknowledgements:**

Support for the Genetic Epidemiology Network of Arteriopathy (GENOA) was provided by the National Heart, Lung and Blood Institute (U01 HL054457, U01 HL054464, U01 HL054481, R01 HL119443, and R01 HL087660) of the National Institutes of Health. DNA extraction for "NHLBI TOPMed: Genetic Epidemiology Network of Arteriopathy" (phs001345) was performed at the Mayo Clinic Genotyping Core, and WGS was performed at the DNA Sequencing and Gene Analysis Center at the University of Washington (3R01HL055673-18S1) and the Broad Institute (HHSN268201500014C). We would like to thank the GENOA participants.

#### GenSalt

The GenSalt study (dbGaP accession phs000784) is a unique NHLBI- sponsored family feeding-study designed to examine the interaction between genes and dietary sodium and potassium intake on BP. A detailed description of the GenSalt study design and participants has been reported previously [9]. Briefly, 3,142 participants from 633 Han families from rural, north China were ascertained through a proband with untreated pre-hypertension or stage-1 hypertension identified from a population-based BP screening. A total of 1,906 GenSalt probands and their siblings, spouses, and offspring were eligible. Among them, 1,818 took part in the TOPMed WGS program and had BP and covariable data available for the current analysis. Three morning BP measurements were obtained according to a standard protocol during each of the 3-days of baseline observation.  All BP readings were measured by trained and certified observers using a random–zero sphygmomanometer using a standard protocol [10]. BP was measured with the participant in the sitting position after 5 minutes of rest. In addition, participants were advised to avoid alcohol, cigarette smoking, coffee/tea, and exercise for at least 30 minutes prior to their BP measurements. Systolic and diastolic BP measures were taken in triplicate during each day of the three-day baseline observation. After throwing out the first measure, the subsequent two measures obtained on the first day of baseline observation were averaged and used in this analysis.

**Ethics statement:**

All subjects provided informed consent and the GenSalt study was approved by the Institutional Review Board (IRB) of all participating institutes in the US and China.

**GenSalt acknowledgements:**

The Genetic Epidemiology Network of Salt-Sensitivity (GenSalt) was supported by research grants (U01HL072507, R01HL087263, and R01HL090682) from the National Heart, Lung and Blood Institute, National Institutes of Health, Bethesda, MD.

#### HCHS/SOL

The Hispanic Community Health Study/Study of Latinos (dbGaP accession phs000810) is a community-based longitudinal cohort study of 16,415 self-identified Hispanic/Latino persons aged 18–74 years and selected from households in predefined census-block groups across four US field centers (in Chicago, Miami, the Bronx, and San Diego). The census-block groups were chosen to provide diversity among cohort participants with regard to socioeconomic status and national origin or background [11, 12]. The HCHS/SOL cohort includes participants who self-identified as having a Hispanic/Latino background; the largest groups are Central American (n = 1,730), Cuban (n = 2,348), Dominican (n = 1,460), Mexican (n = 6,471), Puerto Rican (n = 2,728), and South American (n = 1,068). The HCHS/SOL baseline clinical examination occurred between 2008 and 2011 and included comprehensive biological, behavioral, and sociodemographic assessments. Visit 2 took place between 2014 and 2017, which re-examined 11,623 participants from the baseline sample. Visit 3 has started in 2020 and will last 4 years, ending on January 2024. In addition to clinic visit, participants are contacted annually to assess clinical outcomes. The study was approved by the Institutional Review Boards at each participating institution and written informed consent was obtained from all participants.

**Ethics statement:**

This study was approved by the institutional review boards (IRBs) at each field center, where all participants gave written informed consent, and by the Non-Biomedical IRB at the University of North Carolina at Chapel Hill, to the HCHS/SOL Data Coordinating Center. All IRBs approving the study are: Non-Biomedical IRB at the University of North Carolina at Chapel Hill. Chapel Hill, NC; Einstein IRB at the Albert Einstein College of Medicine of Yeshiva University. Bronx, NY; IRB at Office for the Protection of Research Subjects (OPRS), University of Illinois at Chicago. Chicago,

IL; Human Subject Research Office, University of Miami. Miami, FL; Institutional Review Board of San Diego State University. San Diego, CA.

**HCHS/SOL acknowledgements:**

The Hispanic Community Health Study/Study of Latinos is a collaborative study supported by contracts from the National Heart, Lung, and Blood Institute (NHLBI) to the University of North Carolina (HHSN268201300001I / N01-HC-65233), University of Miami (HHSN268201300004I / N01-HC- 65234), Albert Einstein College of Medicine (HHSN268201300002I / N01-HC-65235), University of Illinois at Chicago – HHSN268201300003I / N01- HC-65236 Northwestern Univ), and San Diego State University (HHSN268201300005I / N01-HC-65237). The following Institutes/Centers/Offices have contributed to the HCHS/SOL through a transfer of funds to the NHLBI: National Institute on Minority Health and Health Disparities, National Institute on Deafness and Other Communication Disorders, National Institute of Dental and Craniofacial Research, National Institute of Diabetes and Digestive and Kidney Diseases, National Institute of Neurological Disorders and Stroke, NIH Institution-Office of Dietary Supplements.

#### JHS

The Jackson Heart Study (dbGaP accession phs000286) is a longitudinal investigation of genetic and environmental risk factors associated with the disproportionate burden of cardiovascular disease in African Americans [13, 14]. At baseline, the JHS recruited 5306 African American residents of the Jackson, Mississippi Metropolitan Statistical Area, which included approximately 6.6% of all African American adults aged 35-84 residing in the area. Participants were recruited via random sampling (17% of participants), volunteers (30%), prior participants in the Atherosclerosis Risk in Communities (ARIC) study (31%), and secondary family members (22%). Among these participants, approximately 3400 gave consent that allows genetic research. JHS participants received three back-to-back clinical examinations (Exam 1, 2000-2004; Exam 2, 2005-2008; and Exam 3, 2009-2013), and a fourth clinical examination started in

2020. Participants are also contacted annually by telephone to update personal and health information including vital status, interim medical events, hospitalizations, functional status and sociocultural information.

**Ethics statement:**

The Institutional Review Boards at Jackson State University, Tougaloo College, and the University of Mississippi Medical Center approved the study, and all participants provided written informed consent.

**JHS acknowledgements:**

The Jackson Heart Study (JHS) is supported and conducted in collaboration with Jackson State University (HHSN268201800013I), Tougaloo College (HHSN268201800014I), the Mississippi State Department of Health (HHSN268201800015I) and the University of Mississippi Medical Center (HHSN268201800010I, HHSN268201800011I and HHSN268201800012I) contracts from the National Heart, Lung, and Blood Institute (NHLBI) and the National Institute for Minority Health and Health Disparities (NIMHD). The authors also wish to thank the staffs and participants of the JHS.

Genome sequencing (dbGap accession phs000964) was performed at the Northwest Genomics Center (HHSN268201100037C). Core support including centralized genomic read mapping and genotype calling, along with variant quality metrics and filtering were provided by the TOPMed Informatics Research Center (R01HL117626; contract HHSN268201800002I). Core support including phenotype harmonization, data management, sample-identity QC, and general program coordination were provided by the TOPMed Data Coordinating Center (R01HL120393; U01HL120393; contract HHSN268201800001I).

#### MESA

The Multi-Ethnic Study of Atherosclerosis (dbGaP accession phs000209) is a study of the characteristics of subclinical cardiovascular disease (disease detected non-invasively before it has produced clinical signs and symptoms) and the risk factors that predict progression to clinically overt cardiovascular disease or progression of the subclinical disease [15]. MESA consisted of a diverse, population-based sample of an initial 6,814 asymptomatic men and women aged 45-84. 38 percent of the recruited participants were white, 28 percent African American, 22 percent Hispanic, and 12 percent Asian, predominantly of Chinese descent. Participants were recruited from six field centers across the United States: Wake Forest University, Columbia University, Johns Hopkins University, University of Minnesota, Northwestern University and University of California - Los Angeles. Participants are being followed for identification and characterization of cardiovascular disease events, including acute myocardial infarction and other forms of coronary heart disease (CHD), stroke, and congestive heart failure; for cardiovascular disease interventions; and for mortality. The first examination took place over two years, from July 2000 - July 2002. It was followed by five examination periods that were 17-20 months in length. Participants have been contacted every 9 to 12 months throughout the study to assess clinical morbidity and mortality.

**Ethics statements:**

All MESA participants provided written informed consent, and the study was approved by the Institutional Review Boards at The Lundquist Institute (formerly Los Angeles BioMedical Research Institute) at Harbor-UCLA Medical Center, University of Washington, Wake Forest School of Medicine, Northwestern University, University of Minnesota, Columbia University, and Johns Hopkins University.

**MESA acknowledgements:**

MESA and the MESA SHARe project are conducted and supported by the National Heart, Lung, and Blood Institute (NHLBI) in collaboration with MESA investigators. Support for MESA is provided by contracts HHSN268201500003I, N01-HC-95159, N01-HC-95160, N01-HC-95161, N01-HC-95162, N01-HC-95163, N01-HC-95164, N01-HC-95165, N01-HC-95166, N01-HC-95167, N01-HC-95168, N01-HC-95169, UL1-TR-000040, UL1-TR-001079, UL1-TR-001420. MESA Family is conducted and supported by the National Heart, Lung, and Blood Institute (NHLBI) in collaboration with MESA investigators. Support is provided by grants and contracts R01HL071051, R01HL071205, R01HL071250, R01HL071251, R01HL071258, R01HL071259, and by the National Center for Research Resources, Grant UL1RR033176. The provision of genotyping data was supported in part by the National Center for Advancing Translational Sciences, CTSI grant UL1TR001881, and the National Institute of Diabetes and Digestive and Kidney Disease Diabetes Research Center (DRC) grant DK063491 to the Southern California Diabetes Endocrinology Research Center.

#### THRV

The THRV-TOPMed study comprises 2,353 Taiwan Chinese participants in three cohorts: The SAPPHIRe (Stanford-Asian Pacific Program in Hypertension and Insulin Resistance) Family cohort of approximately 300 hypertensive sibships (N=1,271) and two hospital-based cohorts, the TSGH (Tri-Service General Hospital) cohort (N=160) and the TCVGH (Taichung Veterans General Hospital) cohort (N=922) that provide population-based controls (unrelated hypertensive or non-hypertensive) matched to SAPPHIRe samples. All three cohorts are based in Taiwan. The 1,271 SAPPHIRe subjects were previously recruited as part of the SAPPHIRe Network of the NHLBI-sponsored Family Blood Pressure Program (FBPP). The SAPPHIRe families were recruited to have two or more hypertensive sibs, with some families having one normotensive/hypotensive sib. The two Hospital-based cohorts (TSGH and TCVGH) both recruited unrelated subjects at the SAPPHIRe field centers/hospitals in Taiwan, that matched with the SAPPHIRe subjects for age, sex, and BMI category. Several metabolic variables associated with blood pressure and insulin resistance were measured in the first 5-year SAPPHIRe funding from the NHLBI (1995-2000). Additional phenotyping through return visits and regular follow ups occurred between 2001 and 2008 which included echocardiographic and multi-detector row CT imaging procedures.

**Ethics statements:**

All THRV participants provided informed consent, and the study was approved by the Institutional Review Board at The Lundquist Institute (formerly Los Angeles BioMedical Research Institute, or LA BioMed) at Harbor-UCLA Medical Center, and at Washington University in St. Louis.

**THRV acknowledgments:**

The Rare Variants for Hypertension in Taiwan Chinese (THRV) is supported by the National Heart, Lung, and Blood Institute (NHLBI) grant (R01HL111249) and its participation in TOPMed is supported by an NHLBI supplement (R01HL111249-04S1). THRV is a collaborative study between Washington University in St. Louis, LA BioMed at Harbor UCLA, University of Texas in Houston, Taichung Veterans General Hospital, Taipei Veterans General Hospital, Tri-Service General Hospital, National Health Research Institutes, National Taiwan University, and Baylor University. THRV is based (substantially) on the parent SAPPHIRe study, along with additional population-based and hospital-based cohorts. SAPPHIRe was supported by NHLBI grants (U01HL54527, U01HL54498) and Taiwan funds, and the other cohorts were supported by Taiwan funds.

#### WHI

The Women's Health Initiative (WHI) cohort. The WHI is a prospective national health study focused on identifying optimal strategies for preventing chronic diseases that are the major causes of death and disability in postmenopausal women. The WHI initially recruited 161,808 women between 1993 and 1997 with the goal of including a socio-demographically diverse population with diversity background groups proportionate to the total minority population of US women aged 50-79 years. The WHI consists of two major parts: a set of randomized Clinical Trials and an Observational Study. The WHI Clinical Trials (CT; N=68,132) includes three overlapping components, each a randomized controlled comparison: the Hormone Therapy Trials (HT), Dietary Modification Trial, and Calcium and Vitamin D Trial. A parallel prospective observational study (OS; N = 93,676) examined biomarkers and risk factors associated with various chronic diseases. While the HT trials ended in the mid-2000s, active follow-up of the WHI-CT and WHI-OS cohorts has continued for over 25 years, with the accumulation of large numbers of diverse clinical outcomes, risk factor measurements, medication use, and many other types of data.

**Ethics statement:**

All WHI participants provided informed consent and the study was approved by the Institutional Review Board (IRB) of the Fred Hutchinson Cancer Research Center.

**WHI acknowledgements:**

The WHI program is funded by the National Heart, Lung, and Blood Institute, National Institutes of Health, U.S. Department of Health and Human Services through contracts 75N92021D00001, 75N92021D00002, 75N92021D00003, 75N92021D00004, 75N92021D00005.

#### Supplementary Note 2: Removal of overlap GWAS

Inverse-variance fixed effects meta-analysis (usually performed by large GWAS meta-analysis efforts) when combining two studies:

Let $\widehat{\beta_1}, \widehat{\beta_2}$ be the effect estimates from study 1 and study 2. Let $\widehat{v_1}, \widehat{v_2}$ be their estimated variances. Let $w_1 = \frac{1}{\widehat{v_1}}, w_2 = \frac{1}{\widehat{v_2}}$, Then:

$$\hat{\beta} = \frac{w_1\widehat{\beta_1} + w_2\widehat{\beta_2}}{w_1 + w_2}$$

And

$$\hat{v} = var(\hat{\beta}) = \frac{w_1^2\widehat{v_1} + w_2^2\widehat{v_2}}{(w_1+w_2)^2} = \frac{w_1+w_2}{(w_1+w_2)^2} = \frac{1}{w_1+w_2}.$$

The same formula straightforwardly extends for an arbitrary number of studies. Further, it can be easily shown that the meta-analytic estimator over $m$ studies will be the same regardless of the order of the meta-analysis, meaning, that one can first meta-analyze statistics from studies 1 and 2, next from studies 3 and 4, and finally meta-analyze the results of these two meta-analyses, and receive the same results as from a meta-analysis of the four studies at the same time.

Therefore, if we have meta-analysis results from $m$ studies, and we can obtain (or compute) the meta-analysis results from a subset of $m_1$ studies, we can "back compute" the results from the $m - m_1$ studies. We also assume that the meta-analysis results from $m_1$ studies are the same as a pooled-summary statistics GWAS results for the same $m_1$ studies (even though likely the results are not *precisely* the same due to different modelling).

Suppose that we had $\hat{\beta}, \hat{v}$ (the meta-analytic estimator of all studies), and $\widehat{\beta_1}, \widehat{v_1}$ estimates from $m_1$studies, a subset of these studies, together. We want to compute the estimates $\widehat{\beta_2}, \widehat{v_2}$

because these are independent of our $m_1$ studies and using them will prevent overfitting when computing polygenic risk models.

Clearly:

$$\widehat{v_2} = \frac{1}{w_2} = \frac{\hat{v}}{1 - w_1\hat{v}} = \frac{\hat{v}}{1 - \hat{v}/\widehat{v_1}}$$

And:

$$\widehat{\beta_2} = \frac{\hat{\beta}(w_1 + w_2) - w_1\widehat{\beta_1}}{w_2}.$$

These can be used to compute p-values of the variants based on the non-overlapping sample of $m_2$ studies from the original meta-analysis.

Using these summary statistics, we selected SNPs and their weights to calculate Polygenic Risk Scores (PRS) and included them in the Machine Learning (ML) models as features to predict the phenotypes.

#### Supplementary Note 3: Sensitivity analysis using PRS-CSx-based global PRS

For comparison, we developed global SBP and DBP PRSs, using PRS-CSx [16]. We used the same GWAS summary statistics used for the main analysis, and coupled BBJ with the UKB East Asian LD reference panel, UKB+ICBP (summary statistics after “removing” the estimated effects of overlapping TOPMed White participants) with the UKB European reference panel, and MVP with the UKB African reference panel. Reference panels are implemented in the PRS-CSx software. We required that SNPs have MAF≥ 0.01 in the source GWAS, and used the overlapping SNPs between the GWAS and the reference panel, as well as other default parameters and the “auto” option, as in previous publication [17] (though note that the GWAS summary statistics are different than the ones used in Kurniansyah et al., 2023). The application of PRS-CSx resulted in three ancestry-specific PRSs for each BP trait. For each BP trait, we constructed the three PRSs in the TOPMed dataset using PRSice but without any clumping and thresholding. The three PRSs were then scaled to have mean 0 and variance, and summed. We next trained the genetic model component of the ensemble using conventional linear regression and non-linear ML (implemented using XGBoost), and compared its performance to that of the main analysis model. Supplementary Table 6 reports the complete performance results as attained PVEs from the genetic models (PVEs of predicting residuals from the baseline model) and ensemble models (PVEs of predicting the raw trait) estimated in cross validation on the training dataset, and from the independent test dataset. Overall, performance of prediction models using a single PRS developed by PRS-CSx showed had PVE in comparison to models trained using PRSs developed using PRSice2.

#### Supplementary Note 4: Descriptions of MGB biobank dataset

Samples, genomic data, and health information were obtained from the Mass General Brigham (MGB) Biobank, a biorepository of consented patient samples at Mass General Brigham.

**DNA samples**

DNA samples are processed from whole blood that was collected as a dedicated research draw or as a clinical discard. Dedicated research samples are aimed to be processed within four hours of collection. Clinical discards are processed 24+ hours after collection. Whole blood is spun to buffy coat with a centrifuge and the buffy coat is stored in a freezer up to several months. The buffy coat is then extracted to DNA. The DNA is then placed in an ultralow freezer (-80oC). Each DNA aliquot contains a minimum of 2 ug of DNA. The concentration varies.

**Genotyping**

Samples have been genotyped using three versions of the biobank SNP array offered by Illumina that is designed to capture the diversity of genetic backgrounds across the globe. The first batch of data was generated on the Multi-Ethnic Genotyping Array (MEGA) array, the first release of this SNP array. The second, third, and fourth batches were generated on the Expanded Multi-Ethnic Genotyping Array (MEGA Ex) array. All remaining data were generated on the Multi-Ethnic Global (MEG) BeadChip.

**Imputation**

Prior to performing imputation, files were converted to VCF format, separated by chromosomes. When multiple probes measured the same genotypes, they were checked for concordance and were set to a missing value if the genotypes did not match. Files were uploaded to the Michigan Imputation Server, and Genotypes were imputed using TOPMed reference panel. Genomic coordinates are provided in GRCh38.

**Quality control**

We performed quality control using PLINK (v2.0. We filtered SNPs with low-quality imputation ($r < 0.5$), with missing call rates > 0.1, HWE p-value less than $1x10^{-6}$ and MAF <1%. We computed principal component (PC) using PLINK: we pruned the genotype data using a window size of 1000 variants, sliding across the genome with a step size of 250 variants at a time, filtering out any SNPs with LD $R^2$>0.1. We used unrelated individuals (3rd degree, identified using PLINK) to compute the loadings for the first 10 PCs.

**PRS construction**

We constructed PRS using PRSice2, using the same SNPs as those based on clumping performed on the TOPMed dataset, and otherwise the same methodology.

**Hypertension status based on Curated Disease Populations**

We used the hypertension outcome from the “curated disease populations” provided by the MGB Biobank team. These phenotypes were developed by the Biobank Portal team using both structured and unstructured electronic medical record (EMR) data and clinical, computational and statistical methods. Natural Language Processing (NLP) was used to extract data from narrative text. Chart reviews by disease experts helped identify features and variables associated with particular phenotypes and were also used to validate results of the algorithms. The process produced robust phenotype algorithms that were evaluated using metrics such as sensitivity, the proportion of true positives correctly identified as such, and positive predictive value (PPV), the proportion of individuals classified as cases by the algorithm [16]. The high throughput phenotyping algorithm is as follows:

1. Create an initial phenotype definition using ICD-9 diagnosis codes.
2. Broaden the definition by determining the most up-to-date features (comorbidities, symptoms, medications) that create a more accurate profile of the phenotype when combined with ICD-9 codes. Features are extracted from online medical literature and knowledge bases via an Automated Feature Extraction Protocol (AFEP).
3. Narrow and refine the definition by determining the features that occur most often in the Biobank data. Extract, code, and rank features contained in clinical narratives with Natural Language Processing (NLP).
4. Create a gold-standard patient set for training the method. Query coded EMR data for the set of patients having at least one ICD-9 code for the phenotype. Apply a statistical sampling algorithm to select a random subset of those patients for full chart review. A clinical expert performs a full chart review to classify the patients as positive or negative for the phenotype.
5. Train a statistical model that incorporates all features in the definition to predict the presence or absence of the phenotype against the gold-standard patient set.
6. Apply the trained model to the entire Biobank Population.

The prevalence of hypertension in the MGB Biobank in general at the time of phenotype development (not restricted to the set of individuals used in our analysis) was 42% and the AUC computed based on the 4 steps above was 0.912.

**Ethics statement**

All Biobank subjects have provided their consent to join the MGB Biobank, which includes agreeing to provide a blood sample linked to the electronic medical record. Subjects also agree to be recontacted by the Partners Biobank staff as needed.

**Acknowledgements**

We thank Mass General Brigham Biobank for providing samples, genomic data, and health information data.

#### Supplementary Note 5: TOPMed and CCDG acknowledgements

Molecular data for the Trans-Omics in Precision Medicine (TOPMed) program was supported by the National Heart, Lung and Blood Institute (NHLBI). Genome sequencing for “NHLBI TOPMed: Genetics of Cardiometabolic Health in the Old Order Amish Study” (phs000956) were performed at the Broad Institute of MIT and Harvard (HHSN268201500014C).

Genome sequencing for “NHLBI TOPMed: Whole Genome Sequencing and Related Phenotypes in the Framingham Heart Study” (phs000974.v4.p3) was performed at the Broad Institute Genomics Platform (3R01HL092577-06S1, 3U54HG003067-12S2). Genome sequencing for the “NHLBI TOPMed: Genetic Epidemiology Network of Arteriopathy (GENOA)” (phs001345.v2.p1) was performed at the Broad Institute Genomics Platform (HHSN268201500014C) and the Northwest Genomics Center (3R01HL055673-18S1). Genome sequencing for “NHLBI TOPMed: The Jackson Heart Study” (phs000964.v1.p1) was performed at the Northwest Genomics Center (HHSN268201100037C). Genome sequencing for the “NHLBI TOPMed: The Atherosclerosis Risk in Communities Study” (phs001211.v3.p2) was performed at the Baylor College of Medicine Human Genome Sequencing Center (HHSN268201500015C and 3U54HG003273-12S2) and the Broad Institute for MIT and Harvard (3R01HL092577- 06S1). Genome sequencing for “NHLBI TOPMed: Coronary Artery Risk Development in Young Adults Study” (phs001612.v1.p1) was performed at the Baylor College of Medicine Human Genome Sequencing Center (HHSN268201600033I). Genome sequencing for “NHLBI TOPMed: Cleveland Family Study” (phs000954.v3.p2) was performed at the Northwest Genomics Center (3R01HL098433-05S1, HHSN268201600032I). Genome sequencing for “NHLBI TOPMed: Genetic Epidemiology of COPD (COPDGene) in the TOPMed Program” (phs000951) was performed at the University of Washington Northwest Genomics Center (3R01 HL089856-08S1) and the Broad Institute of MIT and Harvard (HHSN268201500014C). Genomics sequencing for “NHLBI TOPMed: Cardiovascular Health Study” (phs001368.v2.p1) was performed at the Baylor College of Medicine Human Genome Sequencing Center (3U54HG003273-12S2, HHSN268201500015C, HHSN268201600033I). Genome sequencing for “NHLBI TOPMed: Hispanic Community Health

Study/Study of Latinos" (phs001395.v1.p1) was performed at the Baylor College of Medicine Human Genome Sequencing Center (HHSN268201600033I). Genome sequencing for "NHLBI TOPMed: Women's Health Initiative (WHI)" (phs001237.v2.p1) was performed at the Broad Institute of MIT and Harvard (HHSN268201500014C). Genome sequencing for "NHLBI TOPMed: Multi-Ethnic Study of Atherosclerosis" (phs001416.v2.p1) was performed at Broad Institute Genomics Platform (HHSN268201500014C, 3U54HG003067-13S1). Genome sequencing for "NHLBI TOPMed: Genetic Epidemiology Network of Salt Sensitivity (GenSalt)" (phs001217.v3.p1) was performed at the Baylor College of Medicine Human Genome Sequencing Center (HHSN268201500015C). Genome sequencing for "NHLBI TOPMed: Rare Variants for Hypertension in Taiwan Chinese (THRV)" (phs001387.v3.p1) was performed at the Baylor College of Medicine Human Genome Sequencing Center (3R01HL111249-04S1, HHSN26820150015C). Genome sequencing for "NHLBI TOPMed: Mount Sinai BioMe Biobank (BioMe)" (phs001644.v3.p2) was performed at the Baylor College of Medicine Human Genome Sequencing Center (HHSN268201600033I) and at McDonnell Genome Institute (3UM1HG008853-01S2, HHSN268201600037I). Core support including centralized genomic read mapping and genotype calling, along with variant quality metrics and filtering were provided by the TOPMed Informatics Research Center (3R01HL-117626-02S1; contract HHSN268201800002I). Core support including phenotype harmonization, data management, sample-identity QC, and general program coordination were provided by the TOPMed Data Coordinating Center (R01HL-120393; U01HL-120393; contract HHSN268201800001I). We gratefully acknowledge the studies and participants who provided biological samples and data for TOPMed. The Genome Sequencing Program (GSP) was funded by the National Human

Genome Research Institute (NHGRI), the National Heart, Lung, and Blood Institute (NHLBI), and the National Eye Institute (NEI). The GSP Coordinating Center (U24 HG008956) contributed to cross-program scientific initiatives and provided logistical and general study coordination. The Centers for Common Disease Genomics (CCDG) program was supported by NHGRI and NHLBI, and whole genome sequencing was performed at the Baylor College of Medicine Human Genome Sequencing Center (UM1 HG008898).

#### Supplementary Note 6: TOPMed consortium members

Namiko Abe[1], Gonçalo Abecasis[2], Francois Aguet[3], Christine Albert[4], Laura Almasy[5], Alvaro Alonso[6], Seth Ament[7], Peter Anderson[8], Pramod Anugu[9], Deborah Applebaum-Bowden[10], Kristin Ardlie[3], Dan Arking[11], Donna K Arnett[12], Allison Ashley-Koch[13], Stella Aslibekyan[14], Tim Assimes[15], Paul Auer[16], Dimitrios Avramopoulos[11], Najib Ayas[17], Adithya Balasubramanian[18], John Barnard[19], Kathleen Barnes[20], R. Graham Barr[21], Emily Barron-Casella[11], Lucas Barwick[22], Terri Beaty[11], Gerald Beck[23], Diane Becker[24], Lewis Becker[11], Rebecca Beer[25], Amber Beitelshees[7], Emelia Benjamin[26], Takis Benos[27], Marcos Bezerra[28], Larry Bielak[2], Joshua Bis[29], Thomas Blackwell[2], John Blangero[30], Eric Boerwinkle[31], Donald W. Bowden[32], Russell Bowler[33], Jennifer Brody[8], Ulrich Broeckel[34], Jai Broome[8], Deborah Brown[35], Karen Bunting[1], Esteban Burchard[36], Carlos Bustamante[37], Erin Buth[38], Brian Cade[39], Jonathan Cardwell[40], Vincent Carey[41], Julie Carrier[42], April Carson[43], Cara Carty[44], Richard Casaburi[45], Juan P Casas Romero[46], James Casella[11], Peter Castaldi[47], Mark Chaffin[3], Christy Chang[7], Yi-Cheng Chang[48], Daniel Chasman[49], Sameer Chavan[40], Bo-Juen Chen[1], Wei-Min Chen[50], Yii-Der Ida Chen[51], Michael Cho[41], Seung Hoan Choi[3], Lee-Ming Chuang[52], Mina Chung[53], Ren-Hua Chung[54], Clary Clish[55], Suzy Comhair[56], Matthew Conomos[38], Elaine Cornell[57], Adolfo Correa[58], Carolyn Crandall[45], James Crapo[59], L. Adrienne Cupples[60], Joanne Curran[61], Jeffrey Curtis[62], Brian Custer[63], Coleen Damcott[7], Dawood Darbar[64], Sean David[65], Colleen Davis[8], Michelle Daya[40], Mariza de

Andrade[66], Lisa de las Fuentes[67], Paul de Vries[68], Michael DeBaun[69], Ranjan Deka[70], Dawn DeMeo[41], Scott Devine[7], Huyen Dinh[18], Harsha Doddapaneni[18], Qing Duan[71], Shannon Dugan-Perez[18], Ravi Duggirala[72], Jon Peter Durda[57], Susan K. Dutcher[73], Charles Eaton[74], Lynette Ekunwe[9], Adel El Boueiz[75], Patrick Ellinor[76], Leslie Emery[8], Serpil Erzurum[19], Charles Farber[50], Jesse Farek[18], Tasha Fingerlin[77], Matthew Flickinger[2], Myriam Fornage[31], Nora Franceschini[78], Chris Frazar[8], Mao Fu[7], Stephanie M. Fullerton[8], Lucinda Fulton[79], Stacey Gabriel[3], Weiniu Gan[25], Shanshan Gao[40], Yan Gao[9], Margery Gass[80], Heather Geiger[81], Bruce Gelb[82], Mark Geraci[27], Soren Germer[1], Robert Gerszten[83], Auyon Ghosh[41], Richard Gibbs[18], Chris Gignoux[15], Mark Gladwin[27], David Glahn[84], Stephanie Gogarten[8], Da-Wei Gong[7], Harald Goring[85], Sharon Graw[86], Kathryn J. Gray[87], Daniel Grine[40], Colin Gross[2], C. Charles Gu[79], Yue Guan[7], Xiuqing Guo[51], Namrata Gupta[3], David M. Haas[88], Jeff Haessler[80], Michael Hall[89], Yi Han[18], Patrick Hanly[90], Daniel Harris[91], Nicola L. Hawley[92], Jiang He[93], Ben Heavner[38], Susan Heckbert[94], Ryan Hernandez[36], David Herrington[95], Craig Hersh[96], Bertha Hidalgo[14], James Hixson[31], Brian Hobbs[41], John Hokanson[40], Elliott Hong[7], Karin Hoth[97], Chao (Agnes) Hsiung[98], Jianhong Hu[18], Yi-Jen Hung[99], Haley Huston[100], Chii Min Hwu[101], Marguerite Ryan Irvin[14], Rebecca Jackson[102], Deepti Jain[8], Cashell Jaquish[103], Jill Johnsen[104], Andrew Johnson[25], Craig Johnson[8], Rich Johnston[6], Kimberly Jones[11], Hyun Min Kang[105], Robert Kaplan[106], Sharon Kardia[2], Shannon Kelly[36], Eimear Kenny[82], Michael Kessler[7], Alyna Khan[8], Ziad Khan[18], Wonji Kim[107], John Kimoff[108], Greg Kinney[109], Barbara Konkle[110], Charles Kooperberg[80], Holly Kramer[111], Christoph Lange[112], Ethan Lange[40], Leslie Lange[113], Cathy Laurie[8], Cecelia Laurie[8], Meryl LeBoff[41], Jiwon Lee[41], Sandra Lee[18], Wen-Jane Lee[101], Jonathon LeFaive[2], David Levine[8], Dan Levy[25], Joshua Lewis[7], Xiaohui Li[51], Yun Li[71], Henry Lin[51], Honghuang Lin[114], Xihong Lin[115], Simin Liu[116], Yongmei Liu[117], Yu Liu[118], Ruth J.F. Loos[119], Steven Lubitz[76], Kathryn Lunetta[114], James Luo[25], Ulysses Magalang[120], Michael Mahaney[61], Barry Make[11], Ani Manichaikul[50], Alisa Manning[121], JoAnn Manson[41], Lisa Martin[122], Melissa Marton[81], Susan Mathai[40], Rasika Mathias[11], Susanne May[38], Patrick McArdle[7], Merry-Lynn McDonald[123], Sean McFarland[107], Stephen McGarvey[124], Daniel McGoldrick[125], Caitlin McHugh[38], Becky McNeil[126], Hao Mei[9], James Meigs[127], Vipin Menon[18], Luisa Mestroni[86], Ginger Metcalf[18], Deborah A Meyers[128], Emmanuel Mignot[129], Julie Mikulla[25], Nancy Min[9], Mollie Minear[130], Ryan L Minster[27], Braxton D. Mitchell[7], Matt Moll[47], Zeineen

Momin[18], May E. Montasser[7], Courtney Montgomery[131], Donna Muzny[18], Josyf C Mychaleckyj[50], Girish Nadkarni[82], Rakhi Naik[11], Take Naseri[132], Pradeep Natarajan[3], Sergei Nekhai[133], Sarah C. Nelson[38], Bonnie Neltner[40], Caitlin Nessner[18], Deborah Nickerson[134], Osuji Nkechinyere[18], Kari North[71], Jeff O'Connell[135], Tim O'Connor[7], Heather Ochs-Balcom[136], Geoffrey Okwuonu[18], Allan Pack[137], David T. Paik[138], Nicholette Palmer[139], James Pankow[140], George Papanicolaou[25], Cora Parker[141], Gina Peloso[142], Juan Manuel Peralta[72], Marco Perez[15], James Perry[7], Ulrike Peters[143], Patricia Peyser[2], Lawrence S Phillips[6], Jacob Pleiness[2], Toni Pollin[7], Wendy Post[144], Julia Powers Becker[145], Meher Preethi Boorgula[40], Michael Preuss[82], Bruce Psaty[8], Pankaj Qasba[25], Dandi Qiao[41], Zhaohui Qin[6], Nicholas Rafaels[146], Laura Raffield[147], Mahitha Rajendran[18], Vasan S. Ramachandran[114], D.C. Rao[79], Laura Rasmussen-Torvik[148], Aakrosh Ratan[50], Susan Redline[47], Robert Reed[7], Catherine Reeves[149], Elizabeth Regan[59], Alex Reiner[150], Muagututiâ€˜a Sefuiva Reupena[151], Ken Rice[8], Stephen Rich[50], Rebecca Robillard[152], Nicolas Robine[81], Dan Roden[153], Carolina Roselli[3], Jerome Rotter[154], Ingo Ruczinski[11], Alexi Runnels[81], Pamela Russell[40], Sarah Ruuska[100], Kathleen Ryan[7], Ester Cerdeira Sabino[155], Danish Saleheen[21], Shabnam Salimi[156], Sejal Salvi[18], Steven Salzberg[11], Kevin Sandow[157], Vijay G. Sankaran[158], Jireh Santibanez[18], Karen Schwander[79], David Schwartz[40], Frank Sciurba[27], Christine Seidman[159], Jonathan Seidman[160], FrÃ©dÃ©ric SÃ©riÃ¨s[161], Vivien Sheehan[162], Stephanie L. Sherman[163], Amol Shetty[7], Aniket Shetty[40], Wayne Hui- Heng Sheu[101], M. Benjamin Shoemaker[164], Brian Silver[165], Edwin Silverman[41], Robert Skomro[166], Albert Vernon Smith[167], Jennifer Smith[2], Josh Smith[8], Nicholas Smith[94], Tanja Smith[1], Sylvia Smoller[106], Beverly Snively[168], Michael Snyder[15], Tamar Sofer[41], Nona Sotoodehnia[8], Adrienne M. Stilp[8], Garrett Storm[169], Elizabeth Streeten[7], Jessica Lasky Su[170], Yun Ju Sung[79], Jody Sylvia[41], Adam Szpiro[8], Daniel Taliun[2], Hua Tang[171], Margaret Taub[11], Kent D. Taylor[172], Matthew Taylor[86], Simeon Taylor[7], Marilyn Telen[13], Timothy A. Thornton[8], Machiko Threlkeld[173], Lesley Tinker[174], David Tirschwell[8], Sarah Tishkoff[175], Hemant Tiwari[176], Catherine Tong[177], Russell Tracy[178], Michael Tsai[140], Dhananjay Vaidya[11], David Van Den Berg[179], Peter VandeHaar[2], Scott Vrieze[140], Tarik Walker[40], Robert Wallace[97], Avram Walts[40], Fei Fei Wang[8], Heming Wang[180], Jiongming Wang[167], Karol Watson[45], Jennifer Watt[18], Daniel E. Weeks[27], Joshua Weinstock[105], Bruce Weir[8], Scott T Weiss[181], Lu-Chen Weng[76], Jennifer Wessel[182], Cristen Willer[62], Kayleen Williams[38], L. Keoki Williams[183], Carla Wilson[41], James

Wilson[184], Lara Winterkorn[81], Quenna Wong[8], Joseph Wu[138], Huichun Xu[7], Lisa Yanek[11], Ivana Yang[40], Ketian Yu[2], Seyedeh Maryam Zekavat[3], Yingze Zhang[185], Snow Xueyan Zhao[59], Wei Zhao[186], Xiaofeng Zhu[187], Michael Zody[1], Sebastian Zoellner[2]

1 - New York Genome Center, New York, New York; 2 - University of Michigan, Ann Arbor, Michigan; 3 - Broad Institute, Cambridge, Massachusetts; 4 - Cedars Sinai, Boston, Massachusetts; 5 - Children's Hospital of Philadelphia, University of Pennsylvania, Philadelphia, Pennsylvania; 6 - Emory University, Atlanta, Georgia; 7 - University of Maryland, Baltimore, Maryland; 8 - University of Washington, Seattle, Washington; 9 - University of Mississippi, Jackson, Mississippi; 10 - National Institutes of Health, Bethesda, Maryland; 11 - Johns Hopkins University, Baltimore, Maryland; 12 - University of Kentucky, Lexington, Kentucky; 13 - Duke University, Durham, North Carolina; 14 - University of Alabama, Birmingham, Alabama; 15 - Stanford University, Stanford, California; 16 - Medical College of Wisconsin, Milwaukee, Wisconsin; 17 - Medicine, Providence Health Care, Vancouver; 18 - Baylor College of Medicine Human Genome Sequencing Center, Houston, Texas; 19 - Cleveland Clinic, Cleveland, Ohio; 20 - Tempus, University of Colorado Anschutz Medical Campus, Aurora, Colorado; 21 - Columbia University, New York, New York; 22 - LTRC, The Emmes Corporation, Rockville, Maryland; 23 - Quantitative Health Sciences, Cleveland Clinic, Cleveland, Ohio; 24 - Medicine, Johns Hopkins University, Baltimore, Maryland; 25 - National Heart, Lung, and Blood Institute, National Institutes of Health, Bethesda, Maryland; 26 - Boston University School of Medicine, Boston University, Massachusetts General Hospital, Boston, Massachusetts; 27 - University of Pittsburgh, Pittsburgh, Pennsylvania; 28 - FundaÃ§Ã£o de Hematologia e Hemoterapia de Pernambuco - Hemope, Recife; 29 - Cardiovascular Health Research Unit, Department of Medicine, University of Washington, Seattle, Washington; 30 - Human Genetics, University of Texas Rio Grande Valley School of Medicine, Brownsville, Texas; 31 - University of Texas Health at Houston, Houston, Texas; 32 - Department of Biochemistry, Wake Forest Baptist Health, Winston-Salem, North Carolina; 33 - National Jewish Health, National Jewish Health, Denver, Colorado; 34 - Pediatrics, Medical College of Wisconsin, Milwaukee, Wisconsin; 35 - Pediatrics, University of Texas Health at Houston, Houston, Texas; 36 - University of California, San

Francisco, San Francisco, California; 37 – Biomedical Data Science, Stanford University, Stanford, California; 38 - Biostatistics, University of Washington, Seattle, Washington; 39 - Brigham and Women's Hospital, Brigham & Women's Hospital, Boston, Massachusetts; 40 - University of Colorado at Denver, Denver, Colorado; 41 - Brigham & Women's Hospital, Boston, Massachusetts; 42 - University of Montreal; 43 - Medicine, University of Mississippi, Jackson, Mississippi; 44 - Washington State University, Pullman, Washington; 45 - University of California, Los Angeles, Los Angeles, California; 46 - Brigham & Women's Hospital; 47 - Medicine, Brigham & Women's Hospital, Boston, Massachusetts; 48 - National Taiwan University, Taipei; 49 - Division of Preventive Medicine, Brigham & Women's Hospital, Boston, Massachusetts; 50 - University of Virginia, Charlottesville, Virginia; 51 - Lundquist Institute, Torrance, California; 52 - National Taiwan University Hospital, National Taiwan University, Taipei; 53 - Cleveland Clinic, Cleveland Clinic, Cleveland, Ohio; 54 - National Health Research Institute Taiwan, Miaoli County; 55 - Metabolomics Platform, Broad Institute, Cambridge, Massachusetts; 56 - Immunity and Immunology, Cleveland Clinic, Cleveland, Ohio; 57 - University of Vermont, Burlington, Vermont; 58 - Population Health Science, University of Mississippi, Jackson, Mississippi; 59 - National Jewish Health, Denver, Colorado; 60 - Biostatistics, Boston University, Boston, Massachusetts; 61 - University of Texas Rio Grande Valley School of Medicine, Brownsville, Texas; 62 - Internal Medicine, University of Michigan, Ann Arbor, Michigan; 63 - Vitalant Research Institute, San Francisco, California; 64 - University of Illinois at Chicago, Chicago, Illinois; 65 - University of Chicago, Chicago, Illinois; 66 - Health Quantitative Sciences Research, Mayo Clinic, Rochester, Minnesota; 67 - Department of Medicine, Cardiovascular Division, Washington University in St Louis, St. Louis, Missouri; 68 - Human Genetics Center, Department of Epidemiology, Human Genetics, and Environmental Sciences, University of Texas Health at Houston, Houston, Texas; 69 - Vanderbilt University, Nashville, Tennessee; 70 - University of Cincinnati, Cincinnati, Ohio; 71 - University of North Carolina, Chapel Hill, North Carolina; 72 - University of Texas Rio Grande Valley School of Medicine, Edinburg, Texas; 73 - Genetics, Washington University in St Louis, St Louis, Missouri; 74 - Brown University, Providence, Rhode Island; 75 - Channing Division of Network Medicine, Harvard University, Cambridge, Massachusetts; 76 - Massachusetts General Hospital, Boston,

Massachusetts; 77 - Center for Genes, Environment and Health, National Jewish Health, Denver, Colorado; 78 - Epidemiology, University of North Carolina, Chapel Hill, North Carolina; 79 - Washington University in St Louis, St Louis, Missouri; 80 - Fred Hutchinson Cancer Research Center, Seattle, Washington; 81 - New York Genome Center, New York City, New York; 82 - Icahn School of Medicine at Mount Sinai, New York, New York; 83 - Beth Israel Deaconess Medical Center, Boston, Massachusetts; 84 - Department of Psychiatry, Boston Children's Hospital, Harvard Medical School, Boston, Massachusetts; 85 - University of Texas Rio Grande Valley School of Medicine, San Antonio, Texas; 86 - University of Colorado Anschutz Medical Campus, Aurora, Colorado; 87 - Obstetrics and Gynecology, Mass General Brigham, Boston, Massachusetts; 88 - OB/GYN, Indiana University, Indianapolis, Indiana; 89 - Cardiology, University of Mississippi, Jackson, Mississippi; 90 - Medicine, University of Calgary, Calgary; 91 - Genetics, University of Maryland, Philadelphia, Pennsylvania; 92 - Department of Chronic Disease Epidemiology, Yale University, New Haven, Connecticut; 93 - Tulane University, New Orleans, Louisiana; 94 - Epidemiology, University of Washington, Seattle, Washington; 95 - Wake Forest Baptist Health, Winston-Salem, North Carolina; 96 - Channing Division of Network Medicine, Brigham & Women's Hospital, Boston, Massachusetts; 97 - University of Iowa, Iowa City, Iowa; 98 - Institute of Population Health Sciences, NHRI, National Health Research Institute Taiwan, Miaoli County; 99 - Tri-Service General Hospital National Defense Medical Center; 100 - Blood Works Northwest, Seattle, Washington; 101 - Taichung Veterans General Hospital Taiwan, Taichung City; 102 - Internal Medicine, DIvision of Endocrinology, Diabetes and Metabolism, Oklahoma State University Medical Center, Columbus, Ohio; 103 - NHLBI, National Heart, Lung, and Blood Institute, National Institutes of Health, Bethesda, Maryland; 104 - Research Institute, Blood Works Northwest, Seattle, Washington; 105 - Biostatistics, University of Michigan, Ann Arbor, Michigan; 106 - Albert Einstein College of Medicine, New York, New York; 107 - Harvard University, Cambridge, Massachusetts; 108 - McGill University, MontrÃ©al; 109 - Epidemiology, University of Colorado at Denver, Aurora, Colorado; 110 - Medicine, Blood Works Northwest, Seattle, Washington; 111 - Public Health Sciences, Loyola University, Maywood, Illinois; 112 - Biostats, Harvard School of Public Health, Boston, Massachusetts; 113 - Medicine, University of Colorado at Denver, Aurora, Colorado; 114 - Boston University, Boston,

Massachusetts; 115 - Harvard School of Public Health, Boston, Massachusetts; 116 - Epidemiology and Medicine, Brown University, Providence, Rhode Island; 117 - Cardiology, Duke University, Durham, North Carolina; 118 - Cardiovascular Institute, Stanford University, Stanford, California; 119 - The Charles Bronfman Institute for Personalized Medicine, Icahn School of Medicine at Mount Sinai, New York, New York; 120 - Division of Pulmonary, Critical Care and Sleep Medicine, Ohio State University, Columbus, Ohio; 121 - Broad Institute, Harvard University, Massachusetts General Hospital; 122 - cardiology, George Washington University,Washington, District of Columbia; 123 - University of Alabama at Birmingham, University of Alabama, Birmingham, Alabama; 124 - Epidemiology, Brown University, Providence, Rhode Island; 125 - Genome Sciences, University of Washington, Seattle, Washington; 126 - RTI International; 127 - Medicine, Massachusetts General Hospital, Boston, Massachusetts; 128 - University of Arizona, Tucson, Arizona; 129 - Center For Sleep Sciences and Medicine, Stanford University, Palo Alto, California; 130 - National Institute of Child Health and Human Development, National Institutes of Health, Bethesda, Maryland; 131 - Genes and Human Disease, Oklahoma Medical Research Foundation, Oklahoma City, Oklahoma; 132 - Ministry of Health, Government of Samoa, Apia; 133 - Howard University, Washington, District of Columbia; 134 - Department of Genome Sciences, University of Washington, Seattle, Washington; 135 - University of Maryland, Balitmore, Maryland; 136 - University at Buffalo, Buffalo, New York; 137 - Division of Sleep Medicine/Department of Medicine, University of Pennsylvania, Philadelphia, Pennsylvania; 138 - Stanford Cardiovascular Institute, Stanford University, Stanford, California; 139 - Biochemistry, Wake Forest Baptist Health, Winston-Salem, North Carolina; 140 - University of Minnesota, Minneapolis, Minnesota; 141 - Biostatistics and Epidemiology Division, RTI International, Research Triangle Park, North Carolina; 142 - Department of Biostatistics, Boston University, Boston, Massachusetts; 143 - Fred Hutch and UW, Fred Hutchinson Cancer Research Center, Seattle, Washington; 144 - Cardiology/Medicine, Johns Hopkins University, Baltimore, Maryland; 145 - Medicine, University of Colorado at Denver, Denver, Colorado; 146 - CCPM, University of Colorado at Denver, Denver, Colorado; 147 - Genetics, University of North Carolina, Chapel Hill, North Carolina; 148 - Northwestern University, Chicago, Illinois; 149 - New York Genome Center, New

York Genome Center, New York City, New York; 150 - Fred Hutchinson Cancer Research Center, University of Washington, Seattle, Washington; 151 - Lutia I Puava Ae Mapu I Fagalele, Apia; 152 - Sleep Research Unit, University of Ottawa Institute for Mental Health Research, University of Ottawa, Ottawa; 153 - Medicine, Pharmacology, Biomedicla Informatics, Vanderbilt University, Nashville, Tennessee; 154 - Pediatrics, Lundquist Institute, Torrance, California; 155 - Faculdade de Medicina, Universidade de Sao Paulo, Sao Paulo; 156 - Pathology, University of Maryland, Seattle, Washington; 157 - TGPS, Lundquist Institute, Torrance, California; 158 - Division of Hematology/Oncology, Harvard University, Boston, Massachusetts; 159 - Genetics, Harvard Medical School, Boston, Massachusetts; 160 - Harvard Medical School, Boston, Massachusetts; 161 - UniversitÃ© Laval, Quebec City; 162 - Pediatrics, Emory University, Atlanta, Georgia; 163 - Human Genetics, Emory University, Atlanta, Georgia; 164 - Medicine/Cardiology, Vanderbilt University, Nashville, Tennessee; 165 - UMass Memorial Medical Center, Worcester, Massachusetts; 166 - University of Saskatchewan, Saskatoon; 167 - University of Michigan; 168 - Biostatistical Sciences, Wake Forest Baptist Health, Winston-Salem, North Carolina; 169 - Genomic Cardiology, University of Colorado at Denver, Aurora, Colorado; 170 - Channing Department of Medicine, Brigham & Women's Hospital, Boston, Massachusetts; 171 - Genetics, Stanford University, Stanford, California; 172 - Institute for Translational Genomics and Populations Sciences, Lundquist Institute, Torrance, California; 173 - University of Washington, Department of Genome Sciences, University of Washington, Seattle, Washington; 174 - Cancer Prevention Division of Public Health Sciences, Fred Hutchinson Cancer Research Center, Seattle, Washington; 175 - Genetics, University of Pennsylvania, Philadelphia, Pennsylvania; 176 - Biostatistics, University of Alabama, Birmingham, Alabama; 177 - Department of Biostatistics, University of Washington, Seattle, Washington; 178 - Pathology & Laboratory Medicine, University of Vermont, Burlington, Vermont; 179 - USC Methylation Characterization Center, University of Southern California, University of Southern California, California; 180 - Brigham & Women's Hospital, Mass General Brigham, Boston, Massachusetts; 181 - Channing Division of Network Medicine, Department of Medicine, Brigham & Women's Hospital, Boston, Massachusetts; 182 - Epidemiology, Indiana University, Indianapolis, Indiana; 183 - Henry Ford Health System, Detroit, Michigan; 184 -

Cardiology, Beth Israel Deaconess Medical Center, Cambridge, Massachusetts; 185 - Medicine, University of Pittsburgh, Pittsburgh, Pennsylvania; 186 - Department of Epidemiology, University of Michigan, Ann Arbor, Michigan; 187 - Department of Population and Quantitative Health Sciences, Case Western Reserve University, Cleveland, Ohio
